# Mortality risk between ages 11 and 22 years among young people with neurodisability in England: a national cohort study using linked health and education data

**DOI:** 10.64898/2026.01.12.26343912

**Authors:** Louise Macaulay, Jennifer Saxton, Tamsin Ford, Stuart Logan, Katie Harron, Ruth Gilbert, Ania Zylbersztejn

## Abstract

**Background:** Mortality risk rises from childhood into early adulthood. Young people with neurodisability (neurological conditions causing functional limitations) may be particularly vulnerable during transition from paediatric to adult services.

**Objectives:** To estimate all-cause and cause-specific mortality risk between ages 11 and 22 years in young people with and without neurodisability in England.

**Methods:** In this national cohort study, we used linked health, education, and mortality records from ECHILD to follow pupils aged 11 years in 2008/09-2014/15 from secondary school entry to age 22. Neurodisability was identified from hospital admission records before age 11, likely capturing more severe or complex conditions, and supplemented with education data for intellectual disability and autism. Gender-specific cumulative mortality risk and relative differences were estimated using Kaplan-Meier curves and Cox proportional hazards models adjusted for academic year. Cause-specific cumulative mortality risk was estimated using competing-risks methods.

**Results:** Among 3,601,180 young people, 143,864 (4.0%) had neurodisability. Between ages 11 and 22 years, 5,565 (0.15%) died; 24% had neurodisability. Cumulative mortality risk was 1.6% (95% Confidence Interval [CI] 1.4-1.8%) among females with neurodisability versus 0.14% (95% CI 0.13-0.15%) among peers without neurodisability (Hazard Ratio [HR] 13.9, 95% CI 12.6-15.4). Among males with neurodisability, cumulative risk was 1.3% (95% CI 1.2-1.4%) versus 0.28% (95% CI 0.27-0.29%; HR 5.3, 95% CI 4.9-5.8) among peers. Most deaths in young people with neurodisability were due to medical causes, with cumulative risks of 1.5% (95% CI 1.3-1.6%; HR 25.9, 95% CI 23.0-29.1) in females and 1.1% (95% CI 0.96-1.2%; HR 14.7, 95% CI 13.3-16.3) in males.

**Conclusions:** Young people with neurodisability had higher mortality risk between ages 11 and 22 years than peers, largely from medical causes, with variation by gender and neurodisability subgroup.

## Background

Neurodisability encompasses long-term conditions of the brain and nervous system that cause functional limitations in movement, cognition, communication, hearing, vision, emotion, or behaviour. Conditions include intellectual disability, autism, epilepsy, cerebral palsy, and genetic conditions such as Down syndrome. These conditions frequently co-occur and are associated with complex, long-term health needs, often requiring specialist paediatric care.^1^ Around 1 in 28 primary school children in England have a neurodisability recorded in hospital data, although this is likely an underestimate as not all children require hospital care.^2^

Compared with peers, young people with neurodisability experience poorer outcomes, including higher rates of school absence and exclusion,^3–6^ lower educational attainment,^4–6^ and greater health service use.^5–8^ They also face an increased risk of death. Among young people aged 0-24 years, mortality risk is estimated to be around 3 to over 29 times higher in those with intellectual disability,^9–13^ and 2-7 times higher in those with autism,^9,14,15^ compared with peers without these conditions, with evidence of increased mortality across a range of other neurodisability conditions.^9^

Transition into adulthood is a particularly vulnerable period for young people during which mortality risk rises,^16^ independence increases, and educational settings change, alongside movement from paediatric to adult services (typically occurring between ages 16 and 18 years in the UK, although timing and duration vary across settings).^17^ For young people with neurodisability, this transition is often associated with disruptions in care, reduced continuity, and an increased risk of unmet health needs.^17,18^ Existing studies typically focus on single conditions or broad age ranges, limiting understanding of how mortality risk varies across neurodisability conditions and during this transition period.

We used linked health and education data for the whole of England to investigate gender-specific mortality risk among young people with neurodisability. We characterised neurodisability recorded before secondary school age (<11 years), and estimated cumulative mortality risk, relative differences, and cause-specific mortality between ages 11 and 22 years compared with peers without neurodisability, spanning the transition from paediatric to adult services.

## Methods

All analyses follow a pre-specified protocol.^19^

### Study Design and Data sources

In this descriptive national cohort study,^20^ we used linked education, health and mortality records for young people in England from the Education and Child Health Insights from Linked Data (ECHILD) database. Education records came from the National Pupil Database (NPD), capturing information on pupils in state-funded schools (approximately 93% of all pupils) including pupil characteristics and Special Educational Needs and Disability (SEND) provision (additional educational support for pupils with health, learning, or behavioural needs).^21,22^ Health information came from Hospital Episode Statistics (HES), containing all National Health Service (NHS) funded hospital admissions, with diagnoses and procedures coded using the International Classification of Diseases 10^th^ revision (ICD-10) and the Office of Population Censuses and Surveys Classification of Interventions and Procedures (OPCS). Office for National Statistics (ONS) mortality records, routinely linked to HES by NHS England, provided date and cause of death (coded using ICD-10 from 2001 onwards).^21^ The linked HES-ONS dataset additionally captures in-hospital deaths recorded at discharge, which may include recent deaths undergoing coroner’s inquest and awaiting registration that do not appear in ONS data.^23^ Health and education records were deterministically linked by NHS England using methods described elsewhere, with high linkage rates that increased over time (from 92% in 1990/91 to 99% in 2004/05).^24^

### Study population

We developed a cohort of young people enrolled in state-funded secondary school (mainstream or specialist provision) in the Spring term of Year 7 (the start of secondary school, usually aged 11-12 years) between 2008/2009 and 2014/2015. Young people were excluded if they did not link to a HESID (unique patient identifier in HES required for linkage to ONS mortality data), had an inconsistent date of birth between HES and NPD (suggesting false links or data entry errors), had unknown or missing gender, or died before follow-up began (Figure S1). Follow-up began on 1 February of Year 7 (after the Spring census date) and continued until the earliest of death or 1 March 2020 (before school records were affected by the first COVID-19 lockdown in England), a maximum of 11 years (Figure S2).

### Defining neurodisability

We identified children with neurodisability using hospital and education records before secondary school. Neurodisability was primarily ascertained from hospital admissions data, which typically captures children with more severe or complex conditions.

Children were classified as having neurodisability if they had a relevant diagnosis or procedure recorded during any hospital admission before Year 7, using a published ICD-10 code list developed with clinical input. This included conditions associated with a high likelihood (>50%) of neurological impairment and functional limitation, spanning a broad range of neurological, developmental, and congenital conditions.^2^

Some conditions, particularly those managed predominantly in community settings, may be under-recorded in hospital data.^2^ To improve ascertainment, we supplemented hospital records with education data for intellectual disability and autism, the only conditions with relevant indicators in these data. Children were additionally classified as having intellectual disability or autism if the condition was recorded as a SEND type of need at any point during primary school (Years 1-6).^22^ ICD-10 codes and SEND types of need are provided in Appendix Tables 1 and 2.

**Table 1.** Characteristics of young people with and without neurodisability in the study cohort.

| Characteristic | Total<br>3,601,180 (100%)<sup>a</sup> | No Neurodisability<br>3,457,316 (96%)<sup>a</sup> | Any Neurodisability<br>143,864 (4.0%)<sup>a</sup> |
| --- | --- | --- | --- |
| **Gender** |  |  |  |
| Females | 1,747,730 (49%) | 1,702,771 (49%) | 44,959 (31%) |
| Males | 1,853,450 (51%) | 1,754,545 (51%) | 98,905 (69%) |
| **Ethnic group** |  |  |  |
| White | 2,863,263 (80%) | 2,746,146 (79%) | 117,117 (81%) |
| Asian | 312,257 (8.7%) | 301,952 (8.7%) | 10,305 (7.2%) |
| Black | 167,526 (4.7%) | 160,897 (4.7%) | 6,629 (4.6%) |
| Chinese | 11,305 (0.31%) | 11,005 (0.32%) | 300 (0.21%) |
| Mixed | 150,906 (4.2%) | 145,078 (4.2%) | 5,828 (4.1%) |
| Any other ethnic group | 44,490 (1.2%) | 43,252 (1.3%) | 1,238 (0.86%) |
| Missing | 51,433 (1.4%) | 48,986 (1.4%) | 2,447 (1.7%) |
| **First language** |  |  |  |
| English | 3,128,212 (87%) | 3,000,728 (87%) | 127,484 (89%) |
| Other | 452,810 (13%) | 439,430 (13%) | 13,380 (9%) |
| Missing | 20,158 (0.56%) | 17,158 (0.50%) | 3,000 (2.1%) |
| **Recorded as FSM eligible** |  |  |  |
| No | 2,968,540 (82%)<sup>†</sup> | 2,864,000 (83%)<sup>†</sup> | 104,540 (73%)<sup>†</sup> |
| Yes | 632,630 (18%)<sup>†</sup> | 593,310 (17%)<sup>†</sup> | 39,320 (27%)<sup>†</sup> |
| Missing | * | * | * |
| **IDACI quintile** |  |  |  |
| Q1 (most deprived) | 864,450 (24%) | 826,101 (24%) | 38,349 (27%) |
| Q2 | 727,270 (20%) | 696,219 (20%) | 31,051 (22%) |
| Q3 | 676,030 (19%) | 649,587 (19%) | 26,443 (18%) |
| Q4 | 667,397 (19%) | 643,713 (19%) | 23,684 (16%) |
| Q5 (least deprived) | 648,763 (18%) | 627,473 (18%) | 21,290 (15%) |
| Missing | 17,270 (0.48%) | 14,223 (0.41%) | 3,047 (2.1%) |
| **Year 7 school** |  |  |  |
| Mainstream | 3,537,481 (98%) | 3,434,433 (99%) | 103,048 (72%) |
| Special | 63,699 (1.8%) | 22,883 (0.66%) | 40,816 (28%) |
| **Ever any SEND provision** |  |  |  |
| No | 2,288,591 (64%) | 2,268,191 (66%) | 20,400 (14%) |
| Yes | 1,312,589 (36%) | 1,189,125 (34%) | 123,464 (86%) |
| **Highest SEND provision** |  |  |  |
| SEN support | 1,183,684 (33%) | 1,131,059 (33%) | 52,625 (37%) |
| EHCP | 128,905 (3.6%) | 58,066 (1.7%) | 70,839 (49%) |
| **Chronic condition (excluding neurodisability)** |  |  |  |
| No | 3,006,601 (83%) | 2,933,272 (85%) | 73,329 (51%) |
| Yes | 594,579 (17%) | 524,044 (15%) | 70,535 (49%) |

<sup>a</sup> n (% of all); \* Counts are suppressed where n <10 to protect confidentiality; <sup>†</sup> Counts are rounded to the nearest 10 to protect confidentiality and percentages are calculated from rounded counts. Totals may not sum exactly due to rounding.
EHCP: Education, Health and Care plan; FSM: Free school meals; IDACI: Income Deprivation Affecting Children Index; SEN: Special Educational Needs; SEND: Special Educational Needs and Disabilities.

**Table 2.**
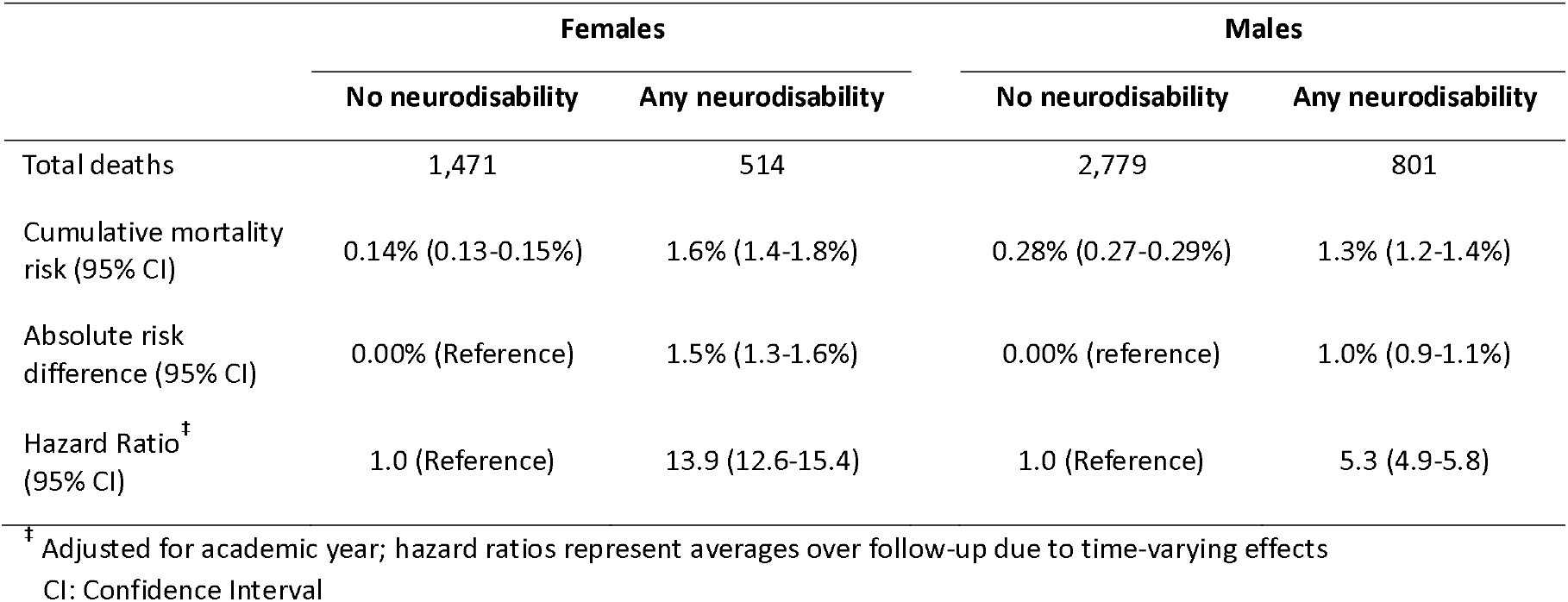
All-cause cumulative mortality risk between ages 11 and 22 years, absolute risk differences, and hazard ratios for young people with and without neurodisability.

|  | Females |  | Males |  |
| --- | --- | --- | --- | --- |
|  | No neurodisability | Any neurodisability | No neurodisability | Any neurodisability |
| Total deaths | 1,471 | 514 | 2,779 | 801 |
| Cumulative mortality risk (95% CI) | 0.14% (0.13-0.15%) | 1.6% (1.4-1.8%) | 0.28% (0.27-0.29%) | 1.3% (1.2-1.4%) |
| Absolute risk difference (95% CI) | 0.00% (Reference) | 1.5% (1.3-1.6%) | 0.00% (reference) | 1.0% (0.9-1.1%) |
| Hazard Ratio<sup>‡</sup> (95% CI) | 1.0 (Reference) | 13.9 (12.6-15.4) | 1.0 (Reference) | 5.3 (4.9-5.8) |

<sup>‡</sup> Adjusted for academic year; hazard ratios represent averages over follow-up due to time-varying effects

#### Neurodisability subgroups

We used the ICD-10 code list and SEND types of need described above to identify 28 neurodisability subgroups, including intellectual disability, autism, cerebral palsy, and epilepsy. Subgroups were not mutually exclusive, as children with neurodisability frequently have multiple co-occurring conditions.^1^

Given under-recording of intellectual disability in hospital data, explicit ICD-10 diagnoses alone capture only a subset of affected children.^25^ We therefore used an expanded code list including: (i) explicit diagnoses of intellectual disability; (ii) “high-risk” conditions (>75% probability of intellectual disability); and (iii) “associated” conditions (30-75% probability), developed through literature review and clinical input, and described elsewhere.^18^

### Cohort characteristics

We extracted pupil characteristics at baseline in Year 7 from the January school census, including gender, ethnic group, first language, school type (mainstream or specialist provision), and two proxy measures of socioeconomic deprivation: Income Deprivation Affecting Children Index (IDACI) quintile and free school meals (FSM) eligibility. Educational needs prior to Year 7 were characterised by any recorded SEND provision and highest level of SEND provision, categorised as special educational needs (SEN) support (school-based additional support) or Education, Health, and Care Plan (EHCP, intensive local government-funded provision for children with complex needs).^22^ We also described any chronic health condition (excluding neurodisability) recorded in a hospital admission prior to Year 7 using a published code list.^26^

### Outcome: mortality

The primary outcome was all-cause mortality, indicated by death recorded at hospital discharge or in ONS death registration. We used a data extract with deaths until June 2021 but stopped follow-up on 1 March 2020, allowing up to 15 months for delayed death registrations.^27^

Our secondary outcome was cause-specific mortality. All recorded causes of death were used to categorise deaths as medical (non-injury deaths with a recorded cause) or injury-related according to ICD-10 codes from a published code list.^28^ Deaths with missing underlying cause information were treated as a separate category and included deaths recorded only in hospital admissions or ONS deaths with no recorded cause.

### Statistical analysis

All analyses were stratified by gender and conducted using R version 4.4.0.

#### All-cause mortality

We described baseline characteristics of pupils in Year 7 and estimated the prevalence of neurodisability prior to Year 7. Using the Kaplan-Meier estimator, we estimated cumulative mortality risk between ages 11 and 22 years for young people with and without neurodisability.^29^ Relative differences in mortality risk were estimated using hazard ratios from Cox proportional hazards regression models,^29^ adjusted for the academic year in which pupils completed Year 7 to account for cohort effects. Additional adjustment was not undertaken, as the aim was to quantify overall differences in mortality between groups.^20^ We assessed proportional hazards assumptions using Schoenfeld residuals. Where violations were identified, time-varying effects were modelled by including interactions between exposure and follow-up time intervals in Cox models, with hazard ratios from models without time interactions interpreted as average effects over follow-up.^30^

#### Cause-specific mortality

We estimated the cumulative mortality risk from medical, injury-related, and missing underlying cause of death using the Aalen-Johansen estimator, accounting for competing risks between causes. Relative risks between young people with and without neurodisability were compared using multi-state Cox proportional hazards models.^29^ We described the distribution of deaths by cause and, for medical causes, by ICD-10 chapter of the underlying cause of death.

#### Neurodisability subgroup mortality

We estimated prevalence and cumulative mortality risk across the 28 neurodisability subgroups to describe the distribution of conditions within the cohort and variation in mortality. We compared subgroup prevalence captured in ECHILD with published population-based data to assess representativeness.

### Sensitivity analyses

Sensitivity analyses assessed the impact of neurodisability case definition on all-cause mortality, comparing: (i) data source for intellectual disability and autism: hospital records alone, education records alone, or both; and (ii) intellectual disability definition, reclassifying cases into mutually exclusive hierarchical tiers of diagnostic certainty: explicit diagnoses, “high-risk” conditions, and “associated” conditions.

### Missing data

Missing data were low for all characteristics (<5%; Table 1). As analyses were descriptive and models were adjusted only for academic year, we did not use multiple imputation. Missing values were retained as a separate category.

### Ethics approval

Ethical approval for analyses of ECHILD were granted by the National Research Ethics Service (17/LO/1494), NHS Health Research Authority Research Ethics Committee (20/EE/0180 and 21/SW/0159), and UCL Great Ormond Street Institute of Child Health’s Joint Research and Development Office (20PE16).

## Results

### Cohort overview

The initial cohort comprised 3,868,256 young people aged 10-12 years enrolled in Year 7. After exclusion of those with inconsistent dates of birth between HES and NPD records (approximately 6,800, 0.18%), those who did not link to a HESID (approximately 260,210, 6.7%), and those who died before follow-up or had missing gender (approximately 70, <0.01%; Figure S1), the final cohort comprised 3,601,180 young people (51% male) with a median follow-up of 8.1 years (interquartile range 6.1-10.1). Compared with the final cohort, young people without a HESID were more likely to be from minority ethnic groups, have a first language other than English, and be less likely to have any SEND provision (Table S1).

Of the final cohort, 4.0% had a record of neurodisability (69% male). Young people with neurodisability were more likely to be male and socioeconomically deprived, and had substantially higher rates of SEND provision, special school attendance, and co-occurring chronic conditions than peers (Table 1).

### All-cause mortality

Between ages 11 and 22 years, 5,565 young people (0.15% of the cohort) had died, of whom nearly one-quarter (24%) had a neurodisability. Among those with neurodisability, around two-thirds (68%) attended special schools and 82% had statutory support plans (EHCPs). Cumulative mortality risk in females with neurodisability was 1.6% (95% Confidence Interval [CI] 1.4-1.8%) compared with 0.14% (95% CI 0.13-0.15%) in females without neurodisability, an absolute difference of 1.5% (95% CI 1.3-1.6%) and a 14 times higher risk (Hazard Ratio [HR] 13.9, 95% CI 12.6-15.4). In males, cumulative risk was 1.3% (95% CI 1.2-1.4%) in those with neurodisability compared with 0.28% (95% CI 0.27-0.29%) in peers, an absolute difference of 1.0% (95% CI 0.9-1.1%) and a 5 times higher risk (HR 5.3, 95% CI 4.9-5.8; Figure 1 and Table 2).

**Figure 1.**
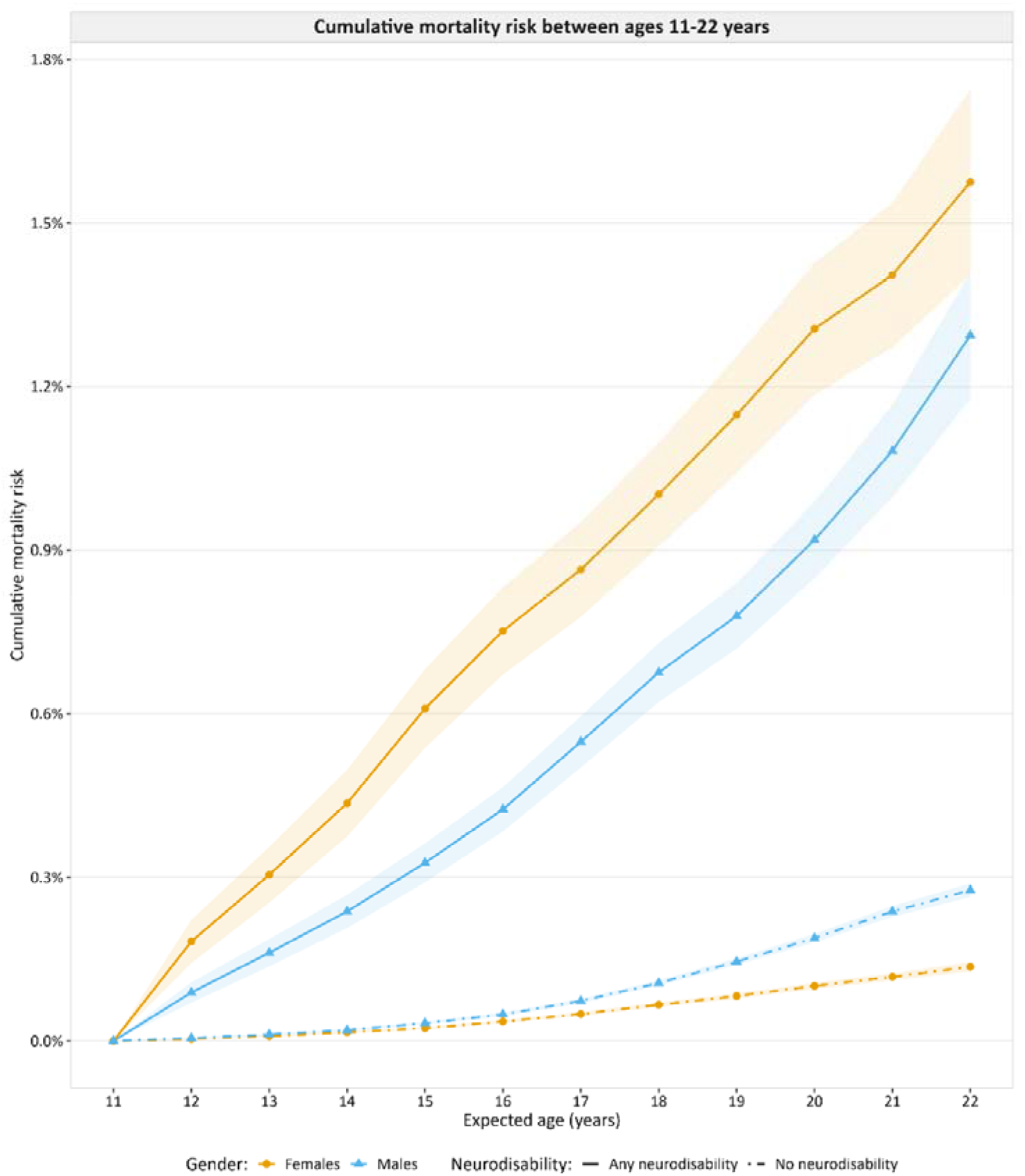
All-cause cumulative mortality risk between ages 11 and 22 years for young people with and without neurodisability. Expected age (years) represents approximate age by school year from the start of Year 7 follow-up (pupils aged 10-12 years at baseline).

The proportional hazards assumption was violated, with hazard ratios highest in early adolescence and generally lower in early adulthood. Hazard ratios were 51.4 (95% CI 36.9-71.6) for females and 16.8 (95% CI 12.6-22.5) for males at ages 11-12 years, compared with 6.6 (95% CI 3.7-11.8) for females at ages 20-21 years and 5.1 (95% CI 3.4-7.6) for males at ages 21-22 years (Figure S3). Therefore, the reported overall hazard ratios represent weighted averages of time-varying effects over the follow-up period.^30^

### Cause-specific mortality

Among young people with neurodisability, most deaths between ages 11 and 22 years were from medical causes (91% in females, 85% in males), compared with less than half in females (49%) and less than one-third in males (31%) without neurodisability. Cumulative mortality risk from medical causes was 1.5% (95% CI 1.3-1.6%) in females with neurodisability compared with 0.07% (95% CI 0.06-0.07%) in peers, corresponding to a nearly 26 times higher risk. In males, cumulative risks were 1.1% (95% CI 0.96-1.2%) in those with neurodisability compared with 0.08% (95% CI 0.07-0.08%) in peers, corresponding to a nearly 15 times higher risk (Table 3). Among medical deaths in young people with neurodisability, the most common underlying causes by ICD-10 chapter were diseases of the nervous system (27% in females, 35% in males), congenital malformations (14% in females, 13% in males), respiratory system diseases (12% in females, 11% in males), and neoplasms (10% in females, 11% in males). By contrast, medical deaths in peers were led by neoplasms (40% in both) and diseases of the circulatory system (14% in females, 19% in males; Table S2).

**Table 3.**
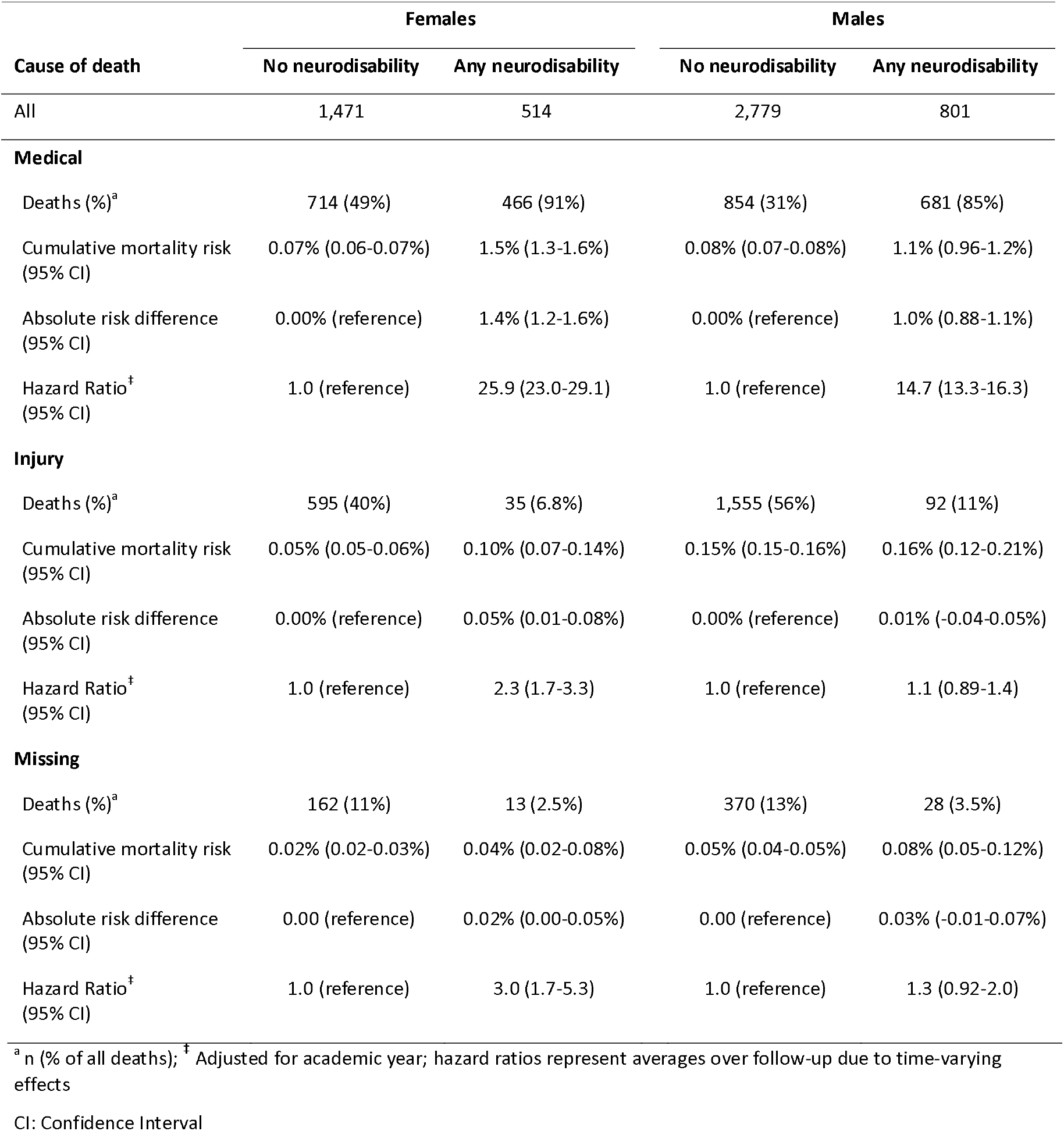
Cause-specific cumulative mortality risk between ages 11 and 22 years, absolute risk differences, and hazard ratios for young people with and without neurodisability.

Injury-related causes accounted for only 6.8% of deaths in females and 11% in males with neurodisability, compared with 40% and 55% in peers, respectively. Cumulative injury-related mortality risk in females with neurodisability (0.10%, 95% CI 0.07-0.14%) was double that of peers (0.05%, 95% CI 0.05-0.06%), while males with and without neurodisability had similar risks (around 0.16%; Table 3).

Cause of death information was more complete for those with neurodisability (missing 2.5% in females, 3.5% in males) than for peers (missing 11% in females, 13% in males). Cumulative risk for deaths with missing underlying cause was low but higher in those with neurodisability (Table 3).

### Neurodisability subgroup mortality

Intellectual disability, including high-risk and associated conditions, was the most prevalent subgroup in females (0.98% in females, 1.6% in males), whereas autism was the most prevalent subgroup in males (0.44% in females, 2.3% in males). Other common subgroups included congenital anomalies (including Down syndrome and central nervous system anomalies), epilepsy, and cerebral palsy. For most subgroups, prevalence was higher in males than females (Figure 2A and Figure ***2***C; Table S3). Prevalence estimates were broadly consistent with published data for conditions typically requiring hospital care, such as cerebral palsy and epilepsy, but were lower than population-based estimates for conditions more commonly identified in community or educational settings, including intellectual disability and autism (Table S4).

**Figure 2.**
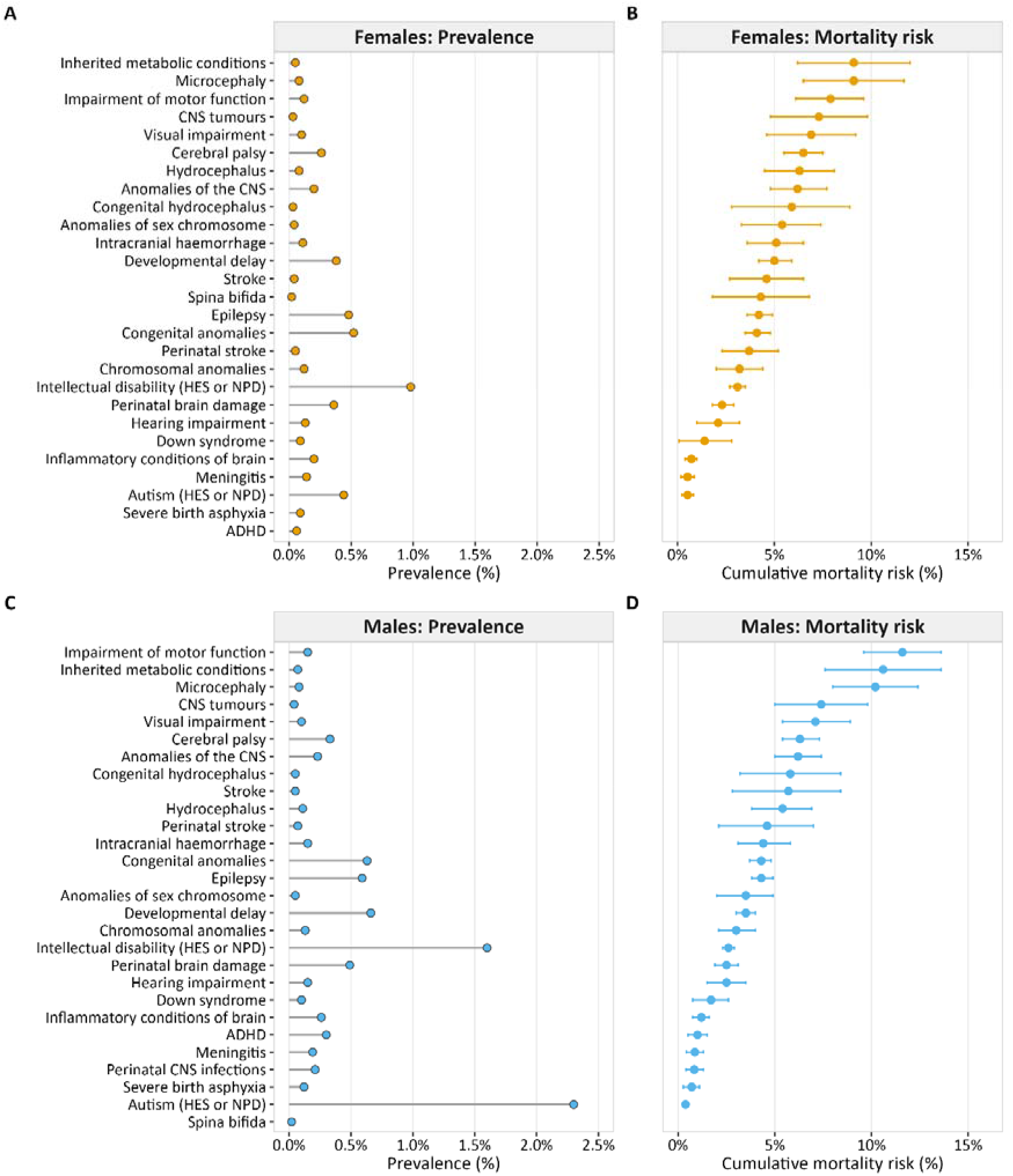
Prevalence and cumulative mortality risk between ages 11 and 22 years for neurodisability subgroups. Mortality risk estimates not calculated where deaths <10. Neurodisability subgroups are non-mutually exclusive. Intellectual disability refers to cases defined using the primary case definition, including explicit diagnoses, high-risk conditions, and associated conditions. All conditions identified from HES data unless otherwise stated. ICD-10 codes and SEND types used to identify conditions are described in Appendix Table 1 and 2. ADHD: Attention Deficit Hyperactivity Disorder; CNS: Central Nervous System; HES: Hospital Episode Statistics; ICD-10: International Classification of Diseases 10th revision; NPD: National Pupil Database.

Cumulative mortality risk between ages 11 and 22 years varied substantially across neurodisability subgroups. Among the described prevalent subgroups, cumulative risk was high among young people with cerebral palsy (6.5%, 95% CI 5.5-7.5% in females; 6.3%, 95% CI 5.4-7.3% in males), epilepsy (4.2%, 95% CI 3.6-4.9% in females; 4.3%, 95% CI 3.8-4.9% in males), congenital anomalies (4.1%, 95% CI 3.5-4.8% in females; 4.3%, 95% CI 3.7-4.8% in males), and intellectual disability (3.1%, 95% CI 2.7-3.5% in females; 2.6%, 95% CI 2.3-2.9% in males), while risk was low among those with autism (0.52%, 95% CI 0.23-0.82% in females; 0.38%, 95% CI 0.29-0.46% in males). Full estimates for all subgroups are presented in Figure 2 and Table S3.

### Sensitivity analyses

Among young people with intellectual disability, cumulative mortality risk was highest in those identified through both hospital and education records (10.1%, 95% CI 8.5-11.6% in females; 9.3%, 95% CI 8.1-10.5% in males), followed by hospital records alone (1.3%, 95% CI 0.88-1.7% in females; 1.6%, 95% CI 1.2-2.0% in males), and education records alone (1.1%, 95% CI 0.81-1.4% in females; 1.1%, 95% CI 0.83-1.3% in males). For autism, cumulative risks were highest in males identified from hospital records alone (1.3%, 95% CI 0.69-1.9%), then both sources (0.51%, 95% CI 0.29-0.73%), and lowest from education records alone (0.28%, 95% CI 0.19-0.37%). Small death counts restricted comparison for females with autism (Table S5).

Among young people with hospital-recorded intellectual disability, 9.1% of females and 11% of males had an explicit diagnosis, 26% of females and 22% of males had a high-risk condition, and 65% of females and 67% of males had an associated condition. Cumulative mortality risk was highest in those with an explicit diagnosis (10.9%, 95% CI 7.7-14.0% in females; 7.3%, 95% CI 5.4-9.1% in males), followed by associated conditions (4.1%, 95% CI 3.3-4.8% in females; 4.7%, 95% CI 4.0-5.3% in males), and lowest in high-risk conditions (3.7%, 95% CI 2.5-5.0% in females; 2.9%, 95% CI 2.1-3.7% in males; Table S6).

### Comment

#### Key findings

In this national cohort of 3.6 million young people in England, 4.0% with neurodisability accounted for nearly one-quarter of all deaths between ages 11 and 22 years. Mortality was substantially higher in young people with neurodisability compared with peers, with larger relative and absolute differences observed among females. Most deaths were due to medical causes, particularly nervous system and respiratory system diseases, congenital malformations, and neoplasms.

#### Strengths

ECHILD’s scale and coverage enabled 11 years of follow-up for 3.6 million young people, with sufficient power to examine rare outcomes and stratify by gender and cause of death. Linkage of health and education records provided context on school type and statutory support, highlighting this population’s complex needs. Estimation of cumulative mortality risk across neurodisability subgroups extends beyond single-condition studies, and stratification by cause and gender highlights distinct mortality patterns that can inform targeted prevention.

#### Limitations

Neurodisability was primarily identified from hospital records, which preferentially capture children with more severe or complex conditions. Conditions typically requiring hospital care were therefore better captured, while those managed predominantly in community settings were under-represented (Table S4). Ascertainment was enhanced using broader code lists and education data for intellectual disability and autism.

We were unable to stratify by condition severity, which is likely to influence mortality risk and should be considered when interpreting subgroup findings. However, sensitivity analyses showed lower mortality risk among young people identified through education records alone, consistent with less complex health needs. Co-occurring conditions may also contribute to observed mortality patterns. Linkage to primary care records would provide more complete coverage of the neurodisability spectrum but is not currently available within ECHILD.

Cause of death data were incomplete, with 10% of deaths lacking a coded underlying cause, likely reflecting delays in death registration, particularly for deaths requiring inquest.^27^ Cause of death recording was more complete for young people with neurodisability than for peers, likely reflecting higher rates of in-hospital deaths. We allowed for a 15-month registration lag to minimise undercounting.

### Interpretation

Mortality risk was elevated among both males and females with neurodisability, with larger relative and absolute differences among females, reversing the usual gender pattern in the general population.^16^ This partly reflects lower baseline mortality in females without neurodisability, but may also reflect differences in case ascertainment or severity. For example, neurodevelopmental conditions such as autism are often under-recognised in females and identified later or only in those with more complex presentations.^31,32^ Diagnostic overshadowing may also contribute to delayed recognition of physical health needs.^33^ Higher hazard ratios at younger ages for both males and females likely reflect lower mortality among peers without neurodisability, although mortality risk remained elevated across all ages.

Mortality varied across neurodisability subgroups. Elevated mortality estimates for young people with conditions such as intellectual disability, cerebral palsy, and epilepsy were broadly consistent with previous population-based studies,^6,9,11,12,34^ while differences across studies, particularly for conditions such as autism,^14,35^ likely reflect variation in case ascertainment and population definitions. For intellectual disability, mortality risk increased with complexity of ascertainment source, consistent with greater clinical need. For autism, risk was highest in males identified from hospital records alone, rather than those identified from both sources, suggesting that educational identification may reflect a less clinically complex group, or that access to educational support may be associated with better outcomes.

Among young people with neurodisability, medical causes dominated, with deaths most commonly from neurological conditions, congenital malformations, respiratory disease, and cancers. Peers died predominantly from injury in males and from a mix of injury and medical causes in females, with medical deaths led by cancers and circulatory disease. The prominence of respiratory disease in the neurodisability population is consistent with evidence that respiratory complications are a leading avoidable cause of death in people with conditions such as intellectual disability and cerebral palsy.^36,37^ These patterns reflect complications of underlying conditions, including seizures, aspiration, and complex co-occurring conditions.^11,12,37–39^ While some deaths reflect progression of severe underlying conditions, others may be amenable to improved recognition of deterioration and better coordination of care.^36,37,39^

These findings are particularly relevant to the transition from paediatric to adult services, often associated with reduced continuity of care.^17^ Young people with neurodisability frequently rely on multiple services, and care fragmentation during transition may increase vulnerability to adverse outcomes. The high level of educational support needs among those who died suggests educational settings may provide important opportunities for coordinated health and education support. Studies including less severe neurodevelopmental conditions are needed to capture the full spectrum of risk, and mechanisms driving gender differences and condition-specific variation warrant further investigation.

## Conclusions

Young people with neurodisability account for a disproportionate share of deaths between ages 11 and 22 years. Mortality differences are driven predominantly by medical causes and are more pronounced among females. These findings highlight the importance of improved continuity of care, proactive monitoring, and coordinated health and education support during transition to adult services, particularly for those with complex health needs. Prevention strategies should be tailored to both gender and condition-specific patterns.

## Data Availability

ECHILD data are not shared publicly in line with data-sharing agreements with NHS Digital and the Department for Education. ECHILD can be accessed by accredited researchers through application via the ECHILD team (www.echild.ac.uk) and the ONS Research Accreditation Panel.

## Author contributions

The study was designed by LM and AZ. LM analysed the data and wrote the first draft of the manuscript. All authors interpreted the data and contributed to subsequent drafts of the manuscript. All authors have seen and approved the final version.

## Acknowledgements

We gratefully acknowledge all children and families whose de-identified data are used in this research. This work was undertaken in the Office for National Statistics (ONS) Secure Research Service using data from ONS and other owners and does not imply the endorsement of the ONS or other data owners.

## Ethics statement

Ethical approval for the ECHILD project was granted by the National Research Ethics Service (17/LO/1494), NHS Health Research Authority Research Ethics Committee (20/EE/0180 and 21/SW/0159) and is overseen by the UCL Great Ormond Street Institute of Child Health’s Joint Research and Development Office (20PE16). Permissions to use linked, de-identified data from HES and the NPD were granted by NHS England (NIC-381972) and DfE (DR200604.02), respectively.

## Consent

Consent from patients is not required here.

## Data Availability Statement

The authors have nothing to report. ECHILD data are not shared publicly in line with data-sharing agreements with NHS Digital and the Department for Education. ECHILD can be accessed by accredited researchers through application via the ECHILD team (www.echild.ac.uk) and the ONS Research Accreditation Panel.

