## appendix for "Mortality risk between ages 11 and 22 years among young people with neurodisability in England: a national cohort study using linked health and education data"

**Appendix Table 1. ICD-10 codes used to classify neurodisability, including subgroup definitions and the expanded intellectual disability classification.**

| **ICD-10 code^†^** | **Description** | **Neurodisability subgroup** ^1^ | **Subtype** | **Intellectual disability classification** ^2^ | **Age criteria**^‡^ |
| --- | --- | --- | --- | --- | --- |
| **Neurodevelopmental conditions** | | | | | |
| F70 | Mild mental retardation | Intellectual disability |  | Explicit diagnosis | 0 |
| F71 | Moderate mental retardation | Intellectual disability |  | Explicit diagnosis | 0 |
| F72 | Severe mental retardation | Intellectual disability |  | Explicit diagnosis | 0 |
| F73 | Profound mental retardation | Intellectual disability |  | Explicit diagnosis | 0 |
| F78 | Other mental retardation | Intellectual disability |  | Explicit diagnosis | 0 |
| F79 | Unspecified mental retardation | Intellectual disability |  | Explicit diagnosis | 0 |
| F80 | Specific developmental disorders of speech and language | Developmental delay |  |  | 0 |
| F800 | Specific speech articulation disorder | Developmental delay |  |  | 0 |
| F801 | Expressive language disorder | Developmental delay |  |  | 0 |
| F802 | Receptive language disorder | Developmental delay |  |  | 0 |
| F808 | Other developmental disorders of speech and language | Developmental delay |  |  | 0 |
| F809 | Developmental disorder of speech and language, unspecified | Developmental delay |  |  | 0 |
| F81 | Specific developmental disorders of scholastic skills | Developmental delay |  |  | 0 |
| F810 | Specific reading disorder | Developmental delay |  |  | 0 |
| F811 | Specific spelling disorder | Developmental delay |  |  | 0 |
| F812 | Specific disorder of arithmetical skills | Developmental delay |  |  | 0 |
| F813 | Mixed disorder of scholastic skills | Developmental delay |  |  | 0 |
| F818 | Other developmental disorders of scholastic skills | Developmental delay |  |  | 0 |
| F819 | Developmental disorder of scholastic skills, unspecified | Developmental delay |  |  | 0 |
| F82 | Specific developmental disorder of motor function | Developmental delay |  |  | 0 |
| F83 | Mixed specific developmental disorders | Developmental delay |  |  | 0 |
| F842 | Rett syndrome | Developmental delay |  | High-risk (>75%) | 0 |
| F88 | Other disorders of psychological development | Developmental delay |  |  | 0 |
| F89 | Unspecified disorder of psychological development | Developmental delay |  |  | 0 |
| F840 | Childhood autism | Autism |  |  | 0 |
| F841 | Atypical autism | Autism |  |  | 0 |
| F843 | Other childhood disintegrative disorder | Autism |  |  | 0 |
| F844 | Overactive disorder associated with mental retardation and stereotyped movements | Autism |  |  | 0 |
| F845 | Asperger syndrome | Autism |  |  | 0 |
| F848 | Other pervasive developmental disorders | Autism |  |  | 0 |
| F849 | Pervasive developmental disorder, unspecified | Autism |  |  | 0 |
| F90 | Hyperkinetic disorders | ADHD |  |  | 0 |
| F900 | Disturbance of activity and attention | ADHD |  |  | 0 |
| F901 | Hyperkinetic conduct disorder | ADHD |  |  | 0 |
| F908 | Other hyperkinetic disorders | ADHD |  |  | 0 |
| F909 | Hyperkinetic disorder, unspecified | ADHD |  |  | 0 |
| F91 | Conduct disorders |  |  |  | 0 |
| F910 | Conduct disorder confined to the family context |  |  |  | 0 |
| F911 | Unsocialized conduct disorder |  |  |  | 0 |
| F912 | Socialized conduct disorder |  |  |  | 0 |
| F913 | Oppositional defiant disorder |  |  |  | 0 |
| F918 | Other conduct disorders |  |  |  | 0 |
| F919 | Conduct disorder, unspecified |  |  |  | 0 |
| F92 | Mixed disorders of conduct and emotions |  |  |  | 0 |
| F920 | Depressive conduct disorder |  |  |  | 0 |
| F928 | Other mixed disorders of conduct and emotions |  |  |  | 0 |
| F929 | Mixed disorder of conduct and emotions, unspecified |  |  |  | 0 |
| F951 | Chronic motor or vocal tic disorder |  |  |  | 0 |
| F952 | Combined vocal and multiple motor tic disorder [de la Tourette] |  |  |  | 0 |
| F958 | Other tic disorders |  |  |  | 0 |
| F959 | Tic disorder, unspecified |  |  |  | 0 |
| F984 | Stereotyped movement disorders |  |  |  | 0 |
| F84 | Pervasive developmental disorders |  |  |  | 0 |
| **Complex neurological conditions** | | | | | |
| G80 | Cerebral palsy | Cerebral palsy |  | Associated (30-75%) | 0 |
| G800 | Spastic quadriplegic cerebral palsy | Cerebral palsy |  | Associated (30-75%) | 0 |
| G801 | Spastic diplegic cerebral palsy | Cerebral palsy |  | Associated (30-75%) | 0 |
| G802 | Spastic hemiplegic cerebral palsy | Cerebral palsy |  | Associated (30-75%) | 0 |
| G803 | Dyskinetic cerebral palsy | Cerebral palsy |  | Associated (30-75%) | 0 |
| G804 | Ataxic cerebral palsy | Cerebral palsy |  | Associated (30-75%) | 0 |
| G808 | Other cerebral palsy | Cerebral palsy |  | Associated (30-75%) | 0 |
| G809 | Cerebral palsy, unspecified | Cerebral palsy |  | Associated (30-75%) | 0 |
| G81 | Hemiplegia | Cerebral palsy |  |  | 0 |
| G810 | Flaccid hemiplegia | Cerebral palsy |  |  | 0 |
| G811 | Spastic hemiplegia | Cerebral palsy |  |  | 0 |
| G819 | Hemiplegia, unspecified | Cerebral palsy |  |  | 0 |
| G82 | Paraplegia and tetraplegia | Cerebral palsy |  |  | 0 |
| G820 | Flaccid paraplegia | Cerebral palsy |  |  | 0 |
| G821 | Spastic paraplegia | Cerebral palsy |  |  | 0 |
| G822 | Paraplegia, unspecified | Cerebral palsy |  |  | 0 |
| G823 | Flaccid tetraplegia | Cerebral palsy |  |  | 0 |
| G824 | Spastic tetraplegia | Cerebral palsy |  |  | 0 |
| G825 | Tetraplegia, unspecified | Cerebral palsy |  |  | 0 |
| G83 | Other paralytic syndromes | Cerebral palsy |  |  | 0 |
| G830 | Diplegia of upper limbs | Cerebral palsy |  |  | 0 |
| G831 | Monoplegia of lower limb | Cerebral palsy |  |  | 0 |
| G832 | Monoplegia of upper limb | Cerebral palsy |  |  | 0 |
| G833 | Monoplegia, unspecified | Cerebral palsy |  |  | 0 |
| F803 | Acquired aphasia with epilepsy [Landau-Kleffner] | Epilepsy |  |  | 0 |
| G40 | Epilepsy | Epilepsy |  |  | 0 |
| G400 | Localization-related (focal)(partial) idiopathic epilepsy and epileptic syndromes with seizures of localized onset | Epilepsy |  |  | 0 |
| G401 | Localization-related (focal)(partial) symptomatic epilepsy and epileptic syndromes with simple partial seizures | Epilepsy |  |  | 0 |
| G402 | Localization-related (focal)(partial) symptomatic epilepsy and epileptic syndromes with complex partial seizures | Epilepsy |  |  | 0 |
| G403 | Generalized idiopathic epilepsy and epileptic syndromes | Epilepsy |  |  | 0 |
| G404 | Other generalized epilepsy and epileptic syndromes | Epilepsy |  |  | 0 |
| G405 | Special epileptic syndromes | Epilepsy |  |  | 0 |
| G406 | Grand mal seizures, unspecified (with or without petit mal) | Epilepsy |  |  | 0 |
| G407 | Petit mal, unspecified, without grand mal seizures | Epilepsy |  |  | 0 |
| G408 | Other epilepsy | Epilepsy |  |  | 0 |
| G409 | Epilepsy, unspecified | Epilepsy |  |  | 0 |
| G41 | Status epilepticus | Epilepsy |  |  | 0 |
| G410 | Grand mal status epilepticus | Epilepsy |  |  | 0 |
| G411 | Petit mal status epilepticus | Epilepsy |  |  | 0 |
| G412 | Complex partial status epilepticus | Epilepsy |  |  | 0 |
| G418 | Other status epilepticus | Epilepsy |  |  | 0 |
| G419 | Status epilepticus, unspecified | Epilepsy |  |  | 0 |
| Y460 | Succinimides | Epilepsy |  |  | 0 |
| Y461 | Oxazolidinediones | Epilepsy |  |  | 0 |
| Y462 | Hydantoin derivatives | Epilepsy |  |  | 0 |
| Y463 | Deoxybarbiturates | Epilepsy |  |  | 0 |
| Y464 | Iminostilbenes | Epilepsy |  |  | 0 |
| Y465 | Valproic acid | Epilepsy |  |  | 0 |
| Y466 | Other and unspecified antiepileptics | Epilepsy |  |  | 0 |
| **Congenital / Chromosomal conditions** | | | | | |
| Q90 | Down syndrome | Chromosomal anomalies | Down syndrome | High-risk (>75%) | 0 |
| Q900 | Trisomy 21, meiotic nondisjunction | Chromosomal anomalies | Down syndrome | High-risk (>75%) | 0 |
| Q901 | Trisomy 21, mosaicism (mitotic nondisjunction) | Chromosomal anomalies | Down syndrome | High-risk (>75%) | 0 |
| Q902 | Trisomy 21, translocation | Chromosomal anomalies | Down syndrome | High-risk (>75%) | 0 |
| Q909 | Down syndrome, unspecified | Chromosomal anomalies | Down syndrome | High-risk (>75%) | 0 |
| Q91 | Edwards' syndrome and Patau syndrome | Chromosomal anomalies |  | High-risk (>75%) | 0 |
| Q910 | Trisomy 18, meiotic nondisjunction | Chromosomal anomalies |  | High-risk (>75%) | 0 |
| Q911 | Trisomy 18, mosaicism (mitotic nondisjunction) | Chromosomal anomalies |  | High-risk (>75%) | 0 |
| Q912 | Trisomy 18, translocation | Chromosomal anomalies |  | High-risk (>75%) | 0 |
| Q913 | Edwards' syndrome, unspecified | Chromosomal anomalies |  | High-risk (>75%) | 0 |
| Q914 | Trisomy 13, meiotic nondisjunction | Chromosomal anomalies |  | High-risk (>75%) | 0 |
| Q915 | Trisomy 13, mosaicism (mitotic nondisjunction) | Chromosomal anomalies |  | High-risk (>75%) | 0 |
| Q916 | Trisomy 13, translocation | Chromosomal anomalies |  | High-risk (>75%) | 0 |
| Q917 | Patau syndrome, unspecified | Chromosomal anomalies |  | High-risk (>75%) | 0 |
| Q92 | Other trisomies and partial trisomies of the autosomes, not elsewhere classified | Chromosomal anomalies |  |  | 0 |
| Q920 | Whole chromosome trisomy, meiotic nondisjunction | Chromosomal anomalies |  | High-risk (>75%) | 0 |
| Q921 | Whole chromosome trisomy, mosaicism (mitotic nondisjunction) | Chromosomal anomalies |  | High-risk (>75%) | 0 |
| Q922 | Major partial trisomy | Chromosomal anomalies |  | High-risk (>75%) | 0 |
| Q923 | Minor partial trisomy | Chromosomal anomalies |  | High-risk (>75%) | 0 |
| Q924 | Duplications seen only at prometaphase | Chromosomal anomalies |  | High-risk (>75%) | 0 |
| Q925 | Duplications with other complex rearrangements | Chromosomal anomalies |  | High-risk (>75%) | 0 |
| Q926 | Extra marker chromosomes | Chromosomal anomalies |  |  | 0 |
| Q927 | Triploidy and polyploidy | Chromosomal anomalies |  | High-risk (>75%) | 0 |
| Q928 | Other specified trisomies and partial trisomies of autosomes | Chromosomal anomalies |  | High-risk (>75%) | 0 |
| Q929 | Trisomy and partial trisomy of autosomes, unspecified | Chromosomal anomalies |  | High-risk (>75%) | 0 |
| Q93 | Monosomies and deletions from the autosomes, not elsewhere classified | Chromosomal anomalies |  | High-risk (>75%) | 0 |
| Q930 | Whole chromosome monosomy, meiotic nondisjunction | Chromosomal anomalies |  | High-risk (>75%) | 0 |
| Q931 | Whole chromosome monosomy, mosaicism (mitotic nondisjunction) | Chromosomal anomalies |  | High-risk (>75%) | 0 |
| Q932 | Chromosome replaced with ring or dicentric | Chromosomal anomalies |  | High-risk (>75%) | 0 |
| Q933 | Deletion of short arm of chromosome 4 | Chromosomal anomalies |  | High-risk (>75%) | 0 |
| Q934 | Deletion of short arm of chromosome 5 | Chromosomal anomalies |  | High-risk (>75%) | 0 |
| Q935 | Other deletions of part of a chromosome | Chromosomal anomalies |  | High-risk (>75%) | 0 |
| Q936 | Deletions seen only at prometaphase | Chromosomal anomalies |  | High-risk (>75%) | 0 |
| Q937 | Deletions with other complex rearrangements | Chromosomal anomalies |  | High-risk (>75%) | 0 |
| Q938 | Other deletions from the autosomes | Chromosomal anomalies |  | High-risk (>75%) | 0 |
| Q939 | Deletion from autosomes, unspecified | Chromosomal anomalies |  | High-risk (>75%) | 0 |
| Q00 | Anencephaly and similar malformations | Anomalies of the CNS |  | High-risk (>75%) | 0 |
| Q000 | Anencephaly | Anomalies of the CNS |  | High-risk (>75%) | 0 |
| Q001 | Craniorachischisis | Anomalies of the CNS |  | High-risk (>75%) | 0 |
| Q002 | Iniencephaly | Anomalies of the CNS |  | High-risk (>75%) | 0 |
| Q01 | Encephalocele | Anomalies of the CNS |  | High-risk (>75%) | 0 |
| Q010 | Frontal encephalocele | Anomalies of the CNS |  | High-risk (>75%) | 0 |
| Q011 | Nasofrontal encephalocele | Anomalies of the CNS |  | High-risk (>75%) | 0 |
| Q012 | Occipital encephalocele | Anomalies of the CNS |  | High-risk (>75%) | 0 |
| Q018 | Encephalocele of other sites | Anomalies of the CNS |  | High-risk (>75%) | 0 |
| Q019 | Encephalocele, unspecified | Anomalies of the CNS |  | High-risk (>75%) | 0 |
| Q02 | Microcephaly | Anomalies of the CNS | Microcephaly | Associated (30-75%) | 0 |
| Q03 | Congenital hydrocephalus | Anomalies of the CNS | Congenital hydrocephalus | Associated (30-75%) | 0 |
| Q030 | Malformations of aqueduct of Sylvius | Anomalies of the CNS | Congenital hydrocephalus | Associated (30-75%) | 0 |
| Q031 | Atresia of foramina of Magendie and Luschka | Anomalies of the CNS | Congenital hydrocephalus | Associated (30-75%) | 0 |
| Q038 | Other congenital hydrocephalus | Anomalies of the CNS | Congenital hydrocephalus | Associated (30-75%) | 0 |
| Q039 | Congenital hydrocephalus, unspecified | Anomalies of the CNS | Congenital hydrocephalus | Associated (30-75%) | 0 |
| Q04 | Other congenital malformations of brain | Anomalies of the CNS |  | Associated (30-75%) | 0 |
| Q040 | Congenital malformations of corpus callosum | Anomalies of the CNS |  | Associated (30-75%) | 0 |
| Q041 | Arhinencephaly | Anomalies of the CNS |  | Associated (30-75%) | 0 |
| Q042 | Holoprosencephaly | Anomalies of the CNS |  | Associated (30-75%) | 0 |
| Q043 | Other reduction deformities of brain | Anomalies of the CNS |  | Associated (30-75%) | 0 |
| Q044 | Septo-optic dysplasia | Anomalies of the CNS |  | Associated (30-75%) | 0 |
| Q045 | Megalencephaly | Anomalies of the CNS |  | Associated (30-75%) | 0 |
| Q046 | Congenital cerebral cysts | Anomalies of the CNS |  | Associated (30-75%) | 0 |
| Q048 | Other specified congenital malformations of brain | Anomalies of the CNS |  | Associated (30-75%) | 0 |
| Q049 | Congenital malformation of brain, unspecified | Anomalies of the CNS |  | Associated (30-75%) | 0 |
| Q05 | Spina bifida | Anomalies of the CNS | Spina bifida |  | 0 |
| Q050 | Cervical spina bifida with hydrocephalus | Anomalies of the CNS | Spina bifida |  | 0 |
| Q051 | Thoracic spina bifida with hydrocephalus | Anomalies of the CNS | Spina bifida |  | 0 |
| Q052 | Lumbar spina bifida with hydrocephalus | Anomalies of the CNS | Spina bifida |  | 0 |
| Q053 | Sacral spina bifida with hydrocephalus | Anomalies of the CNS | Spina bifida |  | 0 |
| Q054 | Unspecified spina bifida with hydrocephalus | Anomalies of the CNS | Spina bifida |  | 0 |
| Q055 | Cervical spina bifida without hydrocephalus | Anomalies of the CNS | Spina bifida |  | 0 |
| Q056 | Thoracic spina bifida without hydrocephalus | Anomalies of the CNS | Spina bifida |  | 0 |
| Q057 | Lumbar spina bifida without hydrocephalus | Anomalies of the CNS | Spina bifida |  | 0 |
| Q058 | Sacral spina bifida without hydrocephalus | Anomalies of the CNS | Spina bifida |  | 0 |
| Q059 | Spina bifida, unspecified | Anomalies of the CNS | Spina bifida |  | 0 |
| Q060 | Amyelia | Anomalies of the CNS |  |  | 0 |
| Q061 | Hypoplasia and dysplasia of spinal cord | Anomalies of the CNS |  |  | 0 |
| Q062 | Diastematomyelia | Anomalies of the CNS |  |  | 0 |
| Q063 | Other congenital cauda equina malformations | Anomalies of the CNS |  |  | 0 |
| Q064 | Hydromyelia | Anomalies of the CNS |  |  | 0 |
| Q068 | Other specified congenital malformations of spinal cord | Anomalies of the CNS |  |  | 0 |
| Q069 | Congenital malformation of spinal cord, unspecified | Anomalies of the CNS |  |  | 0 |
| Q078 | Other specified congenital malformations of nervous system | Anomalies of the CNS |  |  | 0 |
| Q079 | Congenital malformation of nervous system, unspecified | Anomalies of the CNS |  |  | 0 |
| E00 | Congenital iodine-deficiency syndrome |  |  | High-risk (>75%) | 0 |
| E000 | Congenital iodine-deficiency syndrome, neurological type |  |  | High-risk (>75%) | 0 |
| E001 | Congenital iodine-deficiency syndrome, myxoedematous type |  |  | High-risk (>75%) | 0 |
| E002 | Congenital iodine-deficiency syndrome, mixed type |  |  | High-risk (>75%) | 0 |
| E009 | Congenital iodine-deficiency syndrome, unspecified |  |  | High-risk (>75%) | 0 |
| Q860 | Fetal alcohol syndrome (dysmorphic) |  |  | Associated (30-75%) | 0 |
| Q970 | Karyotype 47,XXX | Anomalies of sex chromosome |  |  | 0 |
| Q971 | Female with more than three X chromosomes | Anomalies of sex chromosome |  |  | 0 |
| Q972 | Mosaicism, lines with various numbers of X chromosomes | Anomalies of sex chromosome |  |  | 0 |
| Q978 | Other specified sex chromosome abnormalities, female phenotype | Anomalies of sex chromosome |  |  | 0 |
| Q979 | Sex chromosome abnormality, female phenotype, unspecified | Anomalies of sex chromosome |  |  | 0 |
| Q980 | Klinefelter syndrome karyotype 47,XXY | Anomalies of sex chromosome |  | Associated (30-75%) | 0 |
| Q981 | Klinefelter syndrome, male with more than two X chromosomes | Anomalies of sex chromosome |  | Associated (30-75%) | 0 |
| Q982 | Klinefelter syndrome, male with 46,XX karyotype | Anomalies of sex chromosome |  | Associated (30-75%) | 0 |
| Q983 | Other male with 46,XX karyotype | Anomalies of sex chromosome |  | Associated (30-75%) | 0 |
| Q984 | Klinefelter syndrome, unspecified | Anomalies of sex chromosome |  | Associated (30-75%) | 0 |
| Q985 | Karyotype 47,XYY | Anomalies of sex chromosome |  |  | 0 |
| Q986 | Male with structurally abnormal sex chromosome | Anomalies of sex chromosome |  |  | 0 |
| Q987 | Male with sex chromosome mosaicism | Anomalies of sex chromosome |  |  | 0 |
| Q988 | Other specified sex chromosome abnormalities, male phenotype | Anomalies of sex chromosome |  |  | 0 |
| Q989 | Sex chromosome abnormality, male phenotype, unspecified | Anomalies of sex chromosome |  |  | 0 |
| Q992 | Fragile X chromosome | Anomalies of sex chromosome |  | High-risk (>75%) in males / Associated (30-75%) in females | 0 |
| Q998 | Other specified chromosome abnormalities | Anomalies of sex chromosome |  | High-risk (>75%) | 0 |
| Q999 | Chromosomal abnormality, unspecified | Anomalies of sex chromosome |  | High-risk (>75%) | 0 |
| E702 | Disorders of tyrosine metabolism | Inherited metabolic conditions |  |  | 0 |
| E703 | Albinism | Inherited metabolic conditions |  |  | 0 |
| E708 | Other disorders of aromatic amino-acid metabolism | Inherited metabolic conditions |  |  | 0 |
| E709 | Disorder of aromatic amino-acid metabolism, unspecified | Inherited metabolic conditions |  |  | 0 |
| E71 | Disorders of branched-chain amino-acid metabolism and fatty-acid metabolism | Inherited metabolic conditions |  |  | 0 |
| E710 | Maple-syrup-urine disease | Inherited metabolic conditions |  |  | 0 |
| E711 | Other disorders of branched-chain amino-acid metabolism | Inherited metabolic conditions |  |  | 0 |
| E712 | Disorder of branched-chain amino-acid metabolism, unspecified | Inherited metabolic conditions |  |  | 0 |
| E713 | Disorders of fatty-acid metabolism | Inherited metabolic conditions |  |  | 0 |
| E72 | Other disorders of amino-acid metabolism | Inherited metabolic conditions |  |  | 0 |
| E720 | Disorders of amino-acid transport | Inherited metabolic conditions |  |  | 0 |
| E721 | Disorders of sulfur-bearing amino-acid metabolism | Inherited metabolic conditions |  |  | 0 |
| E722 | Disorders of urea cycle metabolism | Inherited metabolic conditions |  |  | 0 |
| E723 | Disorders of lysine and hydroxylysine metabolism | Inherited metabolic conditions |  |  | 0 |
| E724 | Disorders of ornithine metabolism | Inherited metabolic conditions |  |  | 0 |
| E725 | Disorders of glycine metabolism | Inherited metabolic conditions |  |  | 0 |
| E728 | Other specified disorders of amino-acid metabolism | Inherited metabolic conditions |  |  | 0 |
| E729 | Disorder of amino-acid metabolism, unspecified | Inherited metabolic conditions |  |  | 0 |
| E74 | Other disorders of carbohydrate metabolism | Inherited metabolic conditions |  |  | 0 |
| E740 | Glycogen storage disease | Inherited metabolic conditions |  |  | 0 |
| E741 | Disorders of fructose metabolism | Inherited metabolic conditions |  |  | 0 |
| E742 | Disorders of galactose metabolism | Inherited metabolic conditions |  |  | 0 |
| E743 | Other disorders of intestinal carbohydrate absorption | Inherited metabolic conditions |  |  | 0 |
| E744 | Disorders of pyruvate metabolism and gluconeogenesis | Inherited metabolic conditions |  |  | 0 |
| E748 | Other specified disorders of carbohydrate metabolism | Inherited metabolic conditions |  |  | 0 |
| E749 | Disorder of carbohydrate metabolism, unspecified | Inherited metabolic conditions |  |  | 0 |
| E75 | Disorders of sphingolipid metabolism and other lipid storage disorders | Inherited metabolic conditions |  | High-risk (>75%) | 0 |
| E750 | GM_2_ gangliosidosis | Inherited metabolic conditions |  | High-risk (>75%) | 0 |
| E751 | Other gangliosidosis | Inherited metabolic conditions |  | High-risk (>75%) | 0 |
| E752 | Other sphingolipidosis | Inherited metabolic conditions |  | High-risk (>75%) | 0 |
| E753 | Sphingolipidosis, unspecified | Inherited metabolic conditions |  | High-risk (>75%) | 0 |
| E754 | Neuronal ceroid lipofuscinosis | Inherited metabolic conditions |  | High-risk (>75%) | 0 |
| E755 | Other lipid storage disorders | Inherited metabolic conditions |  | High-risk (>75%) | 0 |
| E756 | Lipid storage disorder, unspecified | Inherited metabolic conditions |  | High-risk (>75%) | 0 |
| E76 | Disorders of glycosaminoglycan metabolism | Inherited metabolic conditions |  | Associated (30-75%) | 0 |
| E760 | Mucopolysaccharidosis, type I | Inherited metabolic conditions |  | Associated (30-75%) | 0 |
| E761 | Mucopolysaccharidosis, type II | Inherited metabolic conditions |  | Associated (30-75%) | 0 |
| E762 | Other mucopolysaccharidoses | Inherited metabolic conditions |  | Associated (30-75%) | 0 |
| E763 | Mucopolysaccharidosis, unspecified | Inherited metabolic conditions |  | Associated (30-75%) | 0 |
| E768 | Other disorders of glucosaminoglycan metabolism | Inherited metabolic conditions |  | Associated (30-75%) | 0 |
| E769 | Disorder of glucosaminoglycan metabolism, unspecified | Inherited metabolic conditions |  | Associated (30-75%) | 0 |
| E77 | Disorders of glycoprotein metabolism | Inherited metabolic conditions |  | Associated (30-75%) | 0 |
| E770 | Defects in post-translational modification of lysosomal enzymes | Inherited metabolic conditions |  | Associated (30-75%) | 0 |
| E771 | Defects in glycoprotein degradation | Inherited metabolic conditions |  | Associated (30-75%) | 0 |
| E778 | Other disorders of glycoprotein metabolism | Inherited metabolic conditions |  | Associated (30-75%) | 0 |
| E779 | Disorder of glycoprotein metabolism, unspecified | Inherited metabolic conditions |  | Associated (30-75%) | 0 |
| E791 | Lesch-Nyhan syndrome | Inherited metabolic conditions |  | High-risk (>75%) | 0 |
| E798 | Other disorders of purine and pyrimidine metabolism | Inherited metabolic conditions |  |  | 0 |
| E799 | Disorder of purine and pyrimidine metabolism, unspecified | Inherited metabolic conditions |  |  | 0 |
| E830 | Disorders of copper metabolism | Inherited metabolic conditions |  | High-risk (>75%) | 0 |
| E851 | Neuropathic heredofamilial amyloidosis | Inherited metabolic conditions |  |  | 0 |
| E888 | Other specified metabolic disorders | Inherited metabolic conditions |  | Associated (30-75%) | 0 |
| E889 | Metabolic disorder, unspecified | Inherited metabolic conditions |  | Associated (30-75%) | 0 |
| D821 | Di George syndrome |  |  |  | 0 |
| Q750 | Craniosynostosis |  |  |  | 0 |
| Q85 | Phakomatoses, not elsewhere classified |  |  |  | 0 |
| Q850 | Neurofibromatosis (nonmalignant) |  |  |  | 0 |
| Q851 | Tuberous sclerosis |  |  | Associated (30-75%) | 0 |
| Q858 | Other phakomatoses, not elsewhere classified |  |  | Associated (30-75%) | 0 |
| Q859 | Phakomatosis, unspecified |  |  | Associated (30-75%) | 0 |
| Q861 | Fetal hydantoin syndrome |  |  | Associated (30-75%) | 0 |
| Q862 | Dysmorphism due to warfarin |  |  | Associated (30-75%) | 0 |
| Q868 | Other congenital malformation syndromes due to known exogenous causes |  |  | Associated (30-75%) | 0 |
| Q870 | Congenital malformation syndromes predominantly affecting facial appearance |  |  | Associated (30-75%) | 0 |
| Q871 | Congenital malformation syndromes predominantly associated with short stature |  |  | Associated (30-75%) | 0 |
| Q872 | Congenital malformation syndromes predominantly involving limbs |  |  | Associated (30-75%) | 0 |
| Q873 | Congenital malformation syndromes involving early overgrowth |  |  | Associated (30-75%) | 0 |
| Q875 | Other congenital malformation syndromes with other skeletal changes |  |  | Associated (30-75%) | 0 |
| Q878 | Other specified congenital malformation syndromes, not elsewhere classified |  |  | Associated (30-75%) | 0 |
| Q899 | Congenital malformation, unspecified |  |  |  | 0 |
| **High-risk conditions affecting the brain** | | | | | |
| G91 | Hydrocephalus | Hydrocephalus |  |  | 0 |
| G910 | Communicating hydrocephalus | Hydrocephalus |  |  | 0 |
| G911 | Obstructive hydrocephalus | Hydrocephalus |  |  | 0 |
| G912 | Normal-pressure hydrocephalus | Hydrocephalus |  |  | 0 |
| G913 | Post-traumatic hydrocephalus, unspecified | Hydrocephalus |  |  | 0 |
| G918 | Other hydrocephalus | Hydrocephalus |  |  | 0 |
| G919 | Hydrocephalus, unspecified | Hydrocephalus |  |  | 0 |
| G940 | Hydrocephalus in infectious and parasitic diseases classified elsewhere | Hydrocephalus |  |  | 0 |
| G941 | Hydrocephalus in neoplastic disease | Hydrocephalus |  |  | 0 |
| G942 | Hydrocephalus in other diseases classified elsewhere | Hydrocephalus |  |  | 0 |
| G972 | Intracranial hypotension following ventricular shunting | Hydrocephalus |  |  | 0 |
| T850 | Mechanical complication of ventricular intracranial (communicating) shunt | Hydrocephalus |  |  | 0 |
| Z982 | Presence of cerebrospinal fluid drainage device | Hydrocephalus |  |  | 0 |
| G46 | Vascular syndromes of brain in cerebrovascular diseases | Stroke |  |  | 0 |
| G460 | Middle cerebral artery syndrome | Stroke |  |  | 0 |
| G461 | Anterior cerebral artery syndrome | Stroke |  |  | 0 |
| G462 | Posterior cerebral artery syndrome | Stroke |  |  | 0 |
| G463 | Brain stem stroke syndrome | Stroke |  |  | 0 |
| G464 | Cerebellar stroke syndrome | Stroke |  |  | 0 |
| G465 | Pure motor lacunar syndrome | Stroke |  |  | 0 |
| G466 | Pure sensory lacunar syndrome | Stroke |  |  | 0 |
| G467 | Other lacunar syndromes | Stroke |  |  | 0 |
| G468 | Other vascular syndromes of brain in cerebrovascular diseases | Stroke |  |  | 0 |
| I60 | Subarachnoid haemorrhage | Stroke |  |  | 0 |
| I600 | Subarachnoid haemorrhage from carotid siphon and bifurcation | Stroke |  |  | 0 |
| I601 | Subarachnoid haemorrhage from middle cerebral artery | Stroke |  |  | 0 |
| I602 | Subarachnoid haemorrhage from anterior communicating artery | Stroke |  |  | 0 |
| I603 | Subarachnoid haemorrhage from posterior communicating artery | Stroke |  |  | 0 |
| I604 | Subarachnoid haemorrhage from basilar artery | Stroke |  |  | 0 |
| I605 | Subarachnoid haemorrhage from vertebral artery | Stroke |  |  | 0 |
| I606 | Subarachnoid haemorrhage from other intracranial arteries | Stroke |  |  | 0 |
| I607 | Subarachnoid haemorrhage from intracranial artery, unspecified | Stroke |  |  | 0 |
| I608 | Other subarachnoid haemorrhage | Stroke |  |  | 0 |
| I609 | Subarachnoid haemorrhage, unspecified | Stroke |  |  | 0 |
| I61 | Intracerebral haemorrhage | Stroke |  |  | 0 |
| I610 | Intracerebral haemorrhage in hemisphere, subcortical | Stroke |  |  | 0 |
| I611 | Intracerebral haemorrhage in hemisphere, cortical | Stroke |  |  | 0 |
| I612 | Intracerebral haemorrhage in hemisphere, unspecified | Stroke |  |  | 0 |
| I613 | Intracerebral haemorrhage in brain stem | Stroke |  |  | 0 |
| I614 | Intracerebral haemorrhage in cerebellum | Stroke |  |  | 0 |
| I615 | Intracerebral haemorrhage, intraventricular | Stroke |  |  | 0 |
| I616 | Intracerebral haemorrhage, multiple localized | Stroke |  |  | 0 |
| I618 | Other intracerebral haemorrhage | Stroke |  |  | 0 |
| I619 | Intracerebral haemorrhage, unspecified | Stroke |  |  | 0 |
| I63 | Cerebral infarction | Stroke |  |  | 0 |
| I630 | Cerebral infarction due to thrombosis of precerebral arteries | Stroke |  |  | 0 |
| I631 | Cerebral infarction due to embolism of precerebral arteries | Stroke |  |  | 0 |
| I632 | Cerebral infarction due to unspecified occlusion or stenosis of precerebral arteries | Stroke |  |  | 0 |
| I633 | Cerebral infarction due to thrombosis of cerebral arteries | Stroke |  |  | 0 |
| I634 | Cerebral infarction due to embolism of cerebral arteries | Stroke |  |  | 0 |
| I635 | Cerebral infarction due to unspecified occlusion or stenosis of cerebral arteries | Stroke |  |  | 0 |
| I636 | Cerebral infarction due to cerebral venous thrombosis, nonpyogenic | Stroke |  |  | 0 |
| I638 | Other cerebral infarction | Stroke |  |  | 0 |
| I639 | Cerebral infarction, unspecified | Stroke |  |  | 0 |
| I64 | Stroke, not specified as haemorrhage or infarction | Stroke |  |  | 0 |
| I67 | Other cerebrovascular diseases | Stroke |  |  | 0 |
| I670 | Dissection of cerebral arteries, nonruptured | Stroke |  |  | 0 |
| I671 | Cerebral aneurysm, nonruptured | Stroke |  |  | 0 |
| I672 | Cerebral atherosclerosis | Stroke |  |  | 0 |
| I673 | Progressive vascular leukoencephalopathy | Stroke |  |  | 0 |
| I674 | Hypertensive encephalopathy | Stroke |  |  | 0 |
| I675 | Moyamoya disease | Stroke |  |  | 0 |
| I676 | Nonpyogenic thrombosis of intracranial venous system | Stroke |  |  | 0 |
| I677 | Cerebral arteritis, not elsewhere classified | Stroke |  |  | 0 |
| I678 | Other specified cerebrovascular diseases | Stroke |  |  | 0 |
| I679 | Cerebrovascular disease, unspecified | Stroke |  |  | 0 |
| I680 | Cerebral amyloid angiopathy | Stroke |  |  | 0 |
| I69 | Sequelae of cerebrovascular disease | Stroke |  |  | 0 |
| I690 | Sequelae of subarachnoid haemorrhage | Stroke |  |  | 0 |
| I691 | Sequelae of intracerebral haemorrhage | Stroke |  |  | 0 |
| I692 | Sequelae of other nontraumatic intracranial haemorrhage | Stroke |  |  | 0 |
| I693 | Sequelae of cerebral infarction | Stroke |  |  | 0 |
| I694 | Sequelae of stroke, not specified as haemorrhage or infarction | Stroke |  |  | 0 |
| I698 | Sequelae of other and unspecified cerebrovascular diseases | Stroke |  |  | 0 |
| I780 | Hereditary haemorrhagic telangiectasia | Stroke |  |  | 0 |
| C70 | Malignant neoplasm of meninges | CNS tumours |  |  | 0 |
| C700 | Malignant neoplasm: Cerebral meninges | CNS tumours |  |  | 0 |
| C701 | Malignant neoplasm: Spinal meninges | CNS tumours |  |  | 0 |
| C709 | Malignant neoplasm: Meninges, unspecified | CNS tumours |  |  | 0 |
| C71 | Malignant neoplasm of brain | CNS tumours |  |  | 0 |
| C710 | Malignant neoplasm: Cerebrum, except lobes and ventricles | CNS tumours |  |  | 0 |
| C711 | Malignant neoplasm: Frontal lobe | CNS tumours |  |  | 0 |
| C712 | Malignant neoplasm: Temporal lobe | CNS tumours |  |  | 0 |
| C713 | Malignant neoplasm: Parietal lobe | CNS tumours |  |  | 0 |
| C714 | Malignant neoplasm: Occipital lobe | CNS tumours |  |  | 0 |
| C715 | Malignant neoplasm: Cerebral ventricle | CNS tumours |  |  | 0 |
| C716 | Malignant neoplasm: Cerebellum | CNS tumours |  |  | 0 |
| C717 | Malignant neoplasm: Brain stem | CNS tumours |  |  | 0 |
| C718 | Malignant neoplasm: Overlapping lesion of brain | CNS tumours |  |  | 0 |
| C719 | Malignant neoplasm: Brain, unspecified | CNS tumours |  |  | 0 |
| C72 | Malignant neoplasm of spinal cord, cranial nerves and other parts of central nervous system | CNS tumours |  |  | 0 |
| C720 | Malignant neoplasm: Spinal cord | CNS tumours |  |  | 0 |
| C721 | Malignant neoplasm: Cauda equina | CNS tumours |  |  | 0 |
| C722 | Malignant neoplasm: Olfactory nerve | CNS tumours |  |  | 0 |
| C723 | Malignant neoplasm: Optic nerve | CNS tumours |  |  | 0 |
| C724 | Malignant neoplasm: Acoustic nerve | CNS tumours |  |  | 0 |
| C725 | Malignant neoplasm: Other and unspecified cranial nerves | CNS tumours |  |  | 0 |
| C728 | Malignant neoplasm: Overlapping lesion of brain and other parts of central nervous system | CNS tumours |  |  | 0 |
| C729 | Malignant neoplasm: Central nervous system, unspecified | CNS tumours |  |  | 0 |
| D32 | Benign neoplasm of meninges | CNS tumours |  |  | 0 |
| D320 | Benign neoplasm: Cerebral meninges | CNS tumours |  |  | 0 |
| D321 | Benign neoplasm: Spinal meninges | CNS tumours |  |  | 0 |
| D329 | Benign neoplasm: Meninges, unspecified | CNS tumours |  |  | 0 |
| D33 | Benign neoplasm of brain and other parts of central nervous system | CNS tumours |  |  | 0 |
| D330 | Benign neoplasm: Brain, supratentorial | CNS tumours |  |  | 0 |
| D331 | Benign neoplasm: Brain, infratentorial | CNS tumours |  |  | 0 |
| D332 | Benign neoplasm: Brain, unspecified | CNS tumours |  |  | 0 |
| D333 | Benign neoplasm: Cranial nerves | CNS tumours |  |  | 0 |
| D334 | Benign neoplasm: Spinal cord | CNS tumours |  |  | 0 |
| D337 | Benign neoplasm: Other specified parts of central nervous system | CNS tumours |  |  | 0 |
| D339 | Benign neoplasm: Central nervous system, unspecified | CNS tumours |  |  | 0 |
| D43 | Neoplasm of uncertain or unknown behaviour of brain and central nervous system | CNS tumours |  |  | 0 |
| D430 | Neoplasm of uncertain or unknown behaviour: Brain, supratentorial | CNS tumours |  |  | 0 |
| D431 | Neoplasm of uncertain or unknown behaviour: Brain, infratentorial | CNS tumours |  |  | 0 |
| D432 | Neoplasm of uncertain or unknown behaviour: Brain, unspecified | CNS tumours |  |  | 0 |
| D433 | Neoplasm of uncertain or unknown behaviour: Cranial nerves | CNS tumours |  |  | 0 |
| D434 | Neoplasm of uncertain or unknown behaviour: Spinal cord | CNS tumours |  |  | 0 |
| D437 | Neoplasm of uncertain or unknown behaviour: Other parts of central nervous system | CNS tumours |  |  | 0 |
| D439 | Neoplasm of uncertain or unknown behaviour: Central nervous system, unspecified | CNS tumours |  |  | 0 |
| A066 | Amoebic brain abscess | Inflammatory conditions of the brain |  |  | 0 |
| A17 | Tuberculosis of nervous system | Inflammatory conditions of the brain |  |  | 0 |
| A170 | Tuberculous meningitis | Inflammatory conditions of the brain | Meningitis |  | 0 |
| A171 | Meningeal tuberculoma | Inflammatory conditions of the brain | Meningitis |  | 0 |
| A178 | Other tuberculosis of nervous system | Inflammatory conditions of the brain |  |  | 0 |
| A179 | Tuberculosis of nervous system, unspecified | Inflammatory conditions of the brain |  |  | 0 |
| A203 | Plague meningitis | Inflammatory conditions of the brain | Meningitis |  | 0 |
| A321 | Listerial meningitis and meningoencephalitis | Inflammatory conditions of the brain | Meningitis |  | 0 |
| A390 | Meningococcal meningitis | Inflammatory conditions of the brain | Meningitis |  | 0 |
| A521 | Symptomatic neurosyphilis | Inflammatory conditions of the brain |  |  | 0 |
| A522 | Asymptomatic neurosyphilis | Inflammatory conditions of the brain |  |  | 0 |
| A523 | Neurosyphilis, unspecified | Inflammatory conditions of the brain |  |  | 0 |
| A800 | Acute paralytic poliomyelitis, vaccine-associated | Inflammatory conditions of the brain |  |  | 0 |
| A801 | Acute paralytic poliomyelitis, wild virus, imported | Inflammatory conditions of the brain |  |  | 0 |
| A802 | Acute paralytic poliomyelitis, wild virus, indigenous | Inflammatory conditions of the brain |  |  | 0 |
| A803 | Acute paralytic poliomyelitis, other and unspecified | Inflammatory conditions of the brain |  |  | 0 |
| A809 | Acute poliomyelitis, unspecified | Inflammatory conditions of the brain |  |  | 0 |
| A82 | Rabies | Inflammatory conditions of the brain |  |  | 0 |
| A820 | Sylvatic rabies | Inflammatory conditions of the brain |  |  | 0 |
| A821 | Urban rabies | Inflammatory conditions of the brain |  |  | 0 |
| A829 | Rabies, unspecified | Inflammatory conditions of the brain |  |  | 0 |
| A83 | Mosquito-borne viral encephalitis | Inflammatory conditions of the brain |  |  | 0 |
| A830 | Japanese encephalitis | Inflammatory conditions of the brain |  |  | 0 |
| A831 | Western equine encephalitis | Inflammatory conditions of the brain |  |  | 0 |
| A832 | Eastern equine encephalitis | Inflammatory conditions of the brain |  |  | 0 |
| A833 | St Louis encephalitis | Inflammatory conditions of the brain |  |  | 0 |
| A834 | Australian encephalitis | Inflammatory conditions of the brain |  |  | 0 |
| A835 | California encephalitis | Inflammatory conditions of the brain |  |  | 0 |
| A836 | Rocio virus disease | Inflammatory conditions of the brain |  |  | 0 |
| A838 | Other mosquito-borne viral encephalitis | Inflammatory conditions of the brain |  |  | 0 |
| A839 | Mosquito-borne viral encephalitis, unspecified | Inflammatory conditions of the brain |  |  | 0 |
| A84 | Tick-borne viral encephalitis | Inflammatory conditions of the brain |  |  | 0 |
| A840 | Far Eastern tick-borne encephalitis [Russian spring-summer encephalitis] | Inflammatory conditions of the brain |  |  | 0 |
| A841 | Central European tick-borne encephalitis | Inflammatory conditions of the brain |  |  | 0 |
| A848 | Other tick-borne viral encephalitis | Inflammatory conditions of the brain |  |  | 0 |
| A849 | Tick-borne viral encephalitis, unspecified | Inflammatory conditions of the brain |  |  | 0 |
| A85 | Other viral encephalitis, not elsewhere classified | Inflammatory conditions of the brain |  |  | 0 |
| A850 | Enteroviral encephalitis | Inflammatory conditions of the brain |  |  | 0 |
| A851 | Adenoviral encephalitis | Inflammatory conditions of the brain |  |  | 0 |
| A852 | Arthropod-borne viral encephalitis, unspecified | Inflammatory conditions of the brain |  |  | 0 |
| A858 | Other specified viral encephalitis | Inflammatory conditions of the brain |  |  | 0 |
| A86 | Unspecified viral encephalitis | Inflammatory conditions of the brain |  |  | 0 |
| B003 | Herpesviral meningitis | Inflammatory conditions of the brain | Meningitis |  | 0 |
| B004 | Herpesviral encephalitis | Inflammatory conditions of the brain |  |  | 0 |
| B010 | Varicella meningitis | Inflammatory conditions of the brain | Meningitis |  | 0 |
| B011 | Varicella encephalitis | Inflammatory conditions of the brain |  |  | 0 |
| B020 | Zoster encephalitis | Inflammatory conditions of the brain |  |  | 0 |
| B021 | Zoster meningitis | Inflammatory conditions of the brain | Meningitis |  | 0 |
| B050 | Measles complicated by encephalitis | Inflammatory conditions of the brain |  |  | 0 |
| B051 | Measles complicated by meningitis | Inflammatory conditions of the brain | Meningitis |  | 0 |
| B060 | Rubella with neurological complications | Inflammatory conditions of the brain |  |  | 0 |
| B261 | Mumps meningitis | Inflammatory conditions of the brain | Meningitis |  | 0 |
| B262 | Mumps encephalitis | Inflammatory conditions of the brain |  |  | 0 |
| B375 | Candidal meningitis | Inflammatory conditions of the brain | Meningitis |  | 0 |
| B384 | Coccidioidomycosis meningitis | Inflammatory conditions of the brain | Meningitis |  | 0 |
| B431 | Phaeomycotic brain abscess | Inflammatory conditions of the brain |  |  | 0 |
| B451 | Cerebral cryptococcosis | Inflammatory conditions of the brain |  |  | 0 |
| B500 | Plasmodium falciparum malaria with cerebral complications | Inflammatory conditions of the brain |  |  | 0 |
| B582 | Toxoplasma meningoencephalitis | Inflammatory conditions of the brain |  |  | 0 |
| B690 | Cysticercosis of central nervous system | Inflammatory conditions of the brain |  |  | 0 |
| B900 | Sequelae of central nervous system tuberculosis | Inflammatory conditions of the brain |  |  | 0 |
| B91 | Sequelae of poliomyelitis | Inflammatory conditions of the brain |  |  | 0 |
| B941 | Sequelae of viral encephalitis | Inflammatory conditions of the brain |  |  | 0 |
| G00 | Bacterial meningitis, not elsewhere classified | Inflammatory conditions of the brain | Meningitis |  | 0 |
| G000 | Haemophilus meningitis | Inflammatory conditions of the brain | Meningitis |  | 0 |
| G001 | Pneumococcal meningitis | Inflammatory conditions of the brain | Meningitis |  | 0 |
| G002 | Streptococcal meningitis | Inflammatory conditions of the brain | Meningitis |  | 0 |
| G003 | Staphylococcal meningitis | Inflammatory conditions of the brain | Meningitis |  | 0 |
| G008 | Other bacterial meningitis | Inflammatory conditions of the brain | Meningitis |  | 0 |
| G009 | Bacterial meningitis, unspecified | Inflammatory conditions of the brain | Meningitis |  | 0 |
| G01 | Meningitis in bacterial diseases classified elsewhere | Inflammatory conditions of the brain | Meningitis |  | 0 |
| G021 | Meningitis in mycoses | Inflammatory conditions of the brain | Meningitis |  | 0 |
| G028 | Meningitis in other specified infectious and parasitic diseases classified elsewhere | Inflammatory conditions of the brain | Meningitis |  | 0 |
| G03 | Meningitis due to other and unspecified causes | Inflammatory conditions of the brain | Meningitis |  | 0 |
| G030 | Nonpyogenic meningitis | Inflammatory conditions of the brain | Meningitis |  | 0 |
| G031 | Chronic meningitis | Inflammatory conditions of the brain | Meningitis |  | 0 |
| G032 | Benign recurrent meningitis [Mollaret] | Inflammatory conditions of the brain | Meningitis |  | 0 |
| G038 | Meningitis due to other specified causes | Inflammatory conditions of the brain | Meningitis |  | 0 |
| G039 | Meningitis, unspecified | Inflammatory conditions of the brain | Meningitis |  | 0 |
| G04 | Encephalitis, myelitis and encephalomyelitis | Inflammatory conditions of the brain |  |  | 0 |
| G040 | Acute disseminated encephalitis | Inflammatory conditions of the brain |  |  | 0 |
| G041 | Tropical spastic paraplegia | Inflammatory conditions of the brain |  |  | 0 |
| G042 | Bacterial meningoencephalitis and meningomyelitis, not elsewhere classified | Inflammatory conditions of the brain |  |  | 0 |
| G048 | Other encephalitis, myelitis and encephalomyelitis | Inflammatory conditions of the brain |  |  | 0 |
| G049 | Encephalitis, myelitis and encephalomyelitis, unspecified | Inflammatory conditions of the brain |  |  | 0 |
| G05 | Encephalitis, myelitis and encephalomyelitis in diseases classified elsewhere | Inflammatory conditions of the brain |  |  | 0 |
| G050 | Encephalitis, myelitis and encephalomyelitis in bacterial diseases classified elsewhere | Inflammatory conditions of the brain |  |  | 0 |
| G051 | Encephalitis, myelitis and encephalomyelitis in viral diseases classified elsewhere | Inflammatory conditions of the brain |  |  | 0 |
| G052 | Encephalitis, myelitis and encephalomyelitis in other infectious and parasitic diseases classified elsewhere | Inflammatory conditions of the brain |  |  | 0 |
| G058 | Encephalitis, myelitis and encephalomyelitis in other diseases classified elsewhere | Inflammatory conditions of the brain |  |  | 0 |
| G06 | Intracranial and intraspinal abscess and granuloma | Inflammatory conditions of the brain |  |  | 0 |
| G060 | Intracranial abscess and granuloma | Inflammatory conditions of the brain |  |  | 0 |
| G061 | Intraspinal abscess and granuloma | Inflammatory conditions of the brain |  |  | 0 |
| G062 | Extradural and subdural abscess, unspecified | Inflammatory conditions of the brain |  |  | 0 |
| G07 | Intracranial and intraspinal abscess and granuloma in diseases classified elsewhere | Inflammatory conditions of the brain |  |  | 0 |
| G08 | Intracranial and intraspinal phlebitis and thrombophlebitis | Inflammatory conditions of the brain |  |  | 0 |
| G09 | Sequelae of inflammatory diseases of central nervous system | Inflammatory conditions of the brain |  |  | 0 |
| M896 | Osteopathy after poliomyelitis | Inflammatory conditions of the brain |  |  | 0 |
| G92 | Toxic encephalopathy |  |  |  | 0 |
| G930 | Cerebral cysts |  |  |  | 0 |
| G931 | Anoxic brain damage, not elsewhere classified |  |  |  | 0 |
| G934 | Encephalopathy, unspecified |  |  |  | 0 |
| G935 | Compression of brain |  |  |  | 0 |
| G936 | Cerebral oedema |  |  |  | 0 |
| G936 | Cerebral oedema |  |  |  | 0 |
| G937 | Reye syndrome |  |  |  | 0 |
| G938 | Other specified disorders of brain |  |  |  | 0 |
| G939 | Disorder of brain, unspecified |  |  |  | 0 |
| G94 | Other disorders of brain in diseases classified elsewhere |  |  |  | 0 |
| G948 | Other specified disorders of brain in diseases classified elsewhere |  |  |  | 0 |
| **Sensory impairments** | | | | | |
| H903 | Sensorineural hearing loss, bilateral | Hearing impairment |  |  | 0 |
| H906 | Mixed conductive and sensorineural hearing loss, bilateral | Hearing impairment |  |  | 0 |
| H913 | Deaf mutism, not elsewhere classified | Hearing impairment |  |  | 0 |
| Z453 | Adjustment and management of implanted hearing device | Hearing impairment |  |  | 0 |
| Z461 | Fitting and adjustment of hearing aid | Hearing impairment |  |  | 0 |
| Z962 | Presence of otological and audiological implants | Hearing impairment |  |  | 0 |
| Z974 | Presence of external hearing-aid | Hearing impairment |  |  | 0 |
| H904 | Sensorineural hearing loss, unilateral with unrestricted hearing on the contralateral side | Hearing impairment |  |  | 0 |
| H905 | Sensorineural hearing loss, unspecified | Hearing impairment |  |  | 0 |
| H907 | Mixed conductive and sensorineural hearing loss, unilateral with unrestricted hearing on the contralateral side | Hearing impairment |  |  | 0 |
| H908 | Mixed conductive and sensorineural hearing loss, unspecified | Hearing impairment |  |  | 0 |
| H540 | Blindness, binocular | Visual impairment |  |  | 0 |
| H541 | Severe visual impairment, binocular | Visual impairment |  |  | 0 |
| H542 | Moderate visual impairment, binocular | Visual impairment |  |  | 0 |
| H351 | Retinopathy of prematurity | Visual impairment |  |  | 0 |
| H185 | Hereditary corneal dystrophies | Visual impairment |  |  | 0 |
| H312 | Hereditary choroidal dystrophy | Visual impairment |  |  | 0 |
| H360 | Diabetic retinopathy | Visual impairment |  |  | 0 |
| H368 | Other retinal disorders in diseases classified elsewhere | Visual impairment |  |  | 0 |
| H472 | Optic atrophy | Visual impairment |  |  | 0 |
| H473 | Other disorders of optic disc | Visual impairment |  |  | 0 |
| H474 | Disorders of optic chiasm | Visual impairment |  |  | 0 |
| H475 | Disorders of other visual pathways | Visual impairment |  |  | 0 |
| H476 | Disorders of visual cortex | Visual impairment |  |  | 0 |
| H477 | Disorder of visual pathways, unspecified | Visual impairment |  |  | 0 |
| H480 | Optic atrophy in diseases classified elsewhere | Visual impairment |  |  | 0 |
| H488 | Other disorders of optic nerve and visual pathways in diseases classified elsewhere | Visual impairment |  |  | 0 |
| H545 | Severe visual impairment, monocular | Visual impairment |  |  | 0 |
| H549 | Unspecified visual impairment (binocular) | Visual impairment |  |  | 0 |
| Q134 | Other congenital corneal malformations | Visual impairment |  |  | 0 |
| Q138 | Other congenital malformations of anterior segment of eye | Visual impairment |  |  | 0 |
| Q139 | Congenital malformation of anterior segment of eye, unspecified | Visual impairment |  |  | 0 |
| Q150 | Congenital glaucoma | Visual impairment |  |  | 0 |
| **Impairment of motor function** | | | | | |
| A810 | Creutzfeldt-Jakob disease |  |  |  | 0 |
| A811 | Subacute sclerosing panencephalitis |  |  |  | 0 |
| A812 | Progressive multifocal leukoencephalopathy |  |  |  | 0 |
| A818 | Other atypical virus infections of central nervous system |  |  |  | 0 |
| G10 | Huntington disease |  |  |  | 0 |
| G11 | Hereditary ataxia |  |  |  | 0 |
| G110 | Congenital nonprogressive ataxia |  |  |  | 0 |
| G111 | Early-onset cerebellar ataxia |  |  |  | 0 |
| G112 | Late-onset cerebellar ataxia |  |  |  | 0 |
| G113 | Cerebellar ataxia with defective DNA repair |  |  |  | 0 |
| G114 | Hereditary spastic paraplegia |  |  |  | 0 |
| G118 | Other hereditary ataxias |  |  |  | 0 |
| G119 | Hereditary ataxia, unspecified |  |  |  | 0 |
| G12 | Spinal muscular atrophy and related syndromes |  |  |  | 0 |
| G120 | Infantile spinal muscular atrophy, type I [Werdnig-Hoffman] |  |  |  | 0 |
| G121 | Other inherited spinal muscular atrophy |  |  |  | 0 |
| G122 | Motor neuron disease |  |  |  | 0 |
| G128 | Other spinal muscular atrophies and related syndromes |  |  |  | 0 |
| G129 | Spinal muscular atrophy, unspecified |  |  |  | 0 |
| G13 | Systemic atrophies primarily affecting central nervous system in diseases classified elsewhere |  |  |  | 0 |
| G130 | Paraneoplastic neuromyopathy and neuropathy |  |  |  | 0 |
| G130 | Paraneoplastic neuromyopathy and neuropathy |  |  |  | 0 |
| G131 | Other systemic atrophy primarily affecting central nervous system in neoplastic disease |  |  |  | 0 |
| G132 | Systemic atrophy primarily affecting central nervous system in myxoedema |  |  |  | 0 |
| G138 | Systemic atrophy primarily affecting central nervous system in other diseases classified elsewhere |  |  |  | 0 |
| G14 | Postpolio syndrome |  |  |  | 0 |
| G35 | Multiple sclerosis |  |  |  | 0 |
| G36 | Other acute disseminated demyelination |  |  |  | 0 |
| G360 | Neuromyelitis optica [Devic] |  |  |  | 0 |
| G361 | Acute and subacute haemorrhagic leukoencephalitis [Hurst] |  |  |  | 0 |
| G368 | Other specified acute disseminated demyelination |  |  |  | 0 |
| G369 | Acute disseminated demyelination, unspecified |  |  |  | 0 |
| G37 | Other demyelinating diseases of central nervous system |  |  |  | 0 |
| G370 | Diffuse sclerosis |  |  |  | 0 |
| G371 | Central demyelination of corpus callosum |  |  |  | 0 |
| G372 | Central pontine myelinolysis |  |  |  | 0 |
| G373 | Acute transverse myelitis in demyelinating disease of central nervous system |  |  |  | 0 |
| G374 | Subacute necrotizing myelitis |  |  |  | 0 |
| G375 | Concentric sclerosis [BalÃ³] |  |  |  | 0 |
| G378 | Other specified demyelinating diseases of central nervous system |  |  |  | 0 |
| G379 | Demyelinating disease of central nervous system, unspecified |  |  |  | 0 |
| G70 | Myasthenia gravis and other myoneural disorders |  |  |  | 0 |
| G700 | Myasthenia gravis |  |  |  | 0 |
| G701 | Toxic myoneural disorders |  |  |  | 0 |
| G702 | Congenital and developmental myasthenia |  |  |  | 0 |
| G708 | Other specified myoneural disorders |  |  |  | 0 |
| G709 | Myoneural disorder, unspecified |  |  |  | 0 |
| G71 | Primary disorders of muscles |  |  |  | 0 |
| G710 | Muscular dystrophy |  |  |  | 0 |
| G711 | Myotonic disorders |  |  |  | 0 |
| G712 | Congenital myopathies |  |  |  | 0 |
| G713 | Mitochondrial myopathy, not elsewhere classified |  |  |  | 0 |
| G718 | Other primary disorders of muscles |  |  |  | 0 |
| G719 | Primary disorder of muscle, unspecified |  |  |  | 0 |
| G723 | Periodic paralysis |  |  |  | 0 |
| G724 | Inflammatory myopathy, not elsewhere classified |  |  |  | 0 |
| G728 | Other specified myopathies |  |  |  | 0 |
| G729 | Myopathy, unspecified |  |  |  | 0 |
| G73 | Disorders of myoneural junction and muscle in diseases classified elsewhere |  |  |  | 0 |
| G730 | Myasthenic syndromes in endocrine diseases |  |  |  | 0 |
| G731 | Lambert-Eaton syndrome |  |  |  | 0 |
| G732 | Other myasthenic syndromes in neoplastic disease |  |  |  | 0 |
| G733 | Myasthenic syndromes in other diseases classified elsewhere |  |  |  | 0 |
| G734 | Myopathy in infectious and parasitic diseases classified elsewhere |  |  |  | 0 |
| G735 | Myopathy in endocrine diseases |  |  |  | 0 |
| G736 | Myopathy in metabolic diseases |  |  |  | 0 |
| G737 | Myopathy in other diseases classified elsewhere |  |  |  | 0 |
| G20 | Parkinson disease |  |  |  | 0 |
| G21 | Secondary parkinsonism |  |  |  | 0 |
| G210 | Malignant neuroleptic syndrome |  |  |  | 0 |
| G211 | Other drug-induced secondary parkinsonism |  |  |  | 0 |
| G212 | Secondary parkinsonism due to other external agents |  |  |  | 0 |
| G213 | Postencephalitic parkinsonism |  |  |  | 0 |
| G214 | Vascular parkinsonism |  |  |  | 0 |
| G218 | Other secondary parkinsonism |  |  |  | 0 |
| G219 | Secondary parkinsonism, unspecified |  |  |  | 0 |
| G22 | Parkinsonism in diseases classified elsewhere |  |  |  | 0 |
| G23 | Other degenerative diseases of basal ganglia |  |  |  | 0 |
| G230 | Hallervorden-Spatz disease |  |  |  | 0 |
| G231 | Progressive supranuclear ophthalmoplegia [Steele-Richardson-Olszewski] |  |  |  | 0 |
| G232 | Multiple system atrophy, parkinsonian type [MSA-P] |  |  |  | 0 |
| G233 | Multiple system atrophy, cerebellar type [MSA-C] |  |  |  | 0 |
| G238 | Other specified degenerative diseases of basal ganglia |  |  |  | 0 |
| G239 | Degenerative disease of basal ganglia, unspecified |  |  |  | 0 |
| G240 | Drug-induced dystonia |  |  |  | 0 |
| G241 | Idiopathic familial dystonia |  |  |  | 0 |
| G242 | Idiopathic nonfamilial dystonia |  |  |  | 0 |
| G243 | Spasmodic torticollis |  |  |  | 0 |
| G244 | Idiopathic orofacial dystonia |  |  |  | 0 |
| G248 | Other dystonia |  |  |  | 0 |
| G249 | Dystonia, unspecified |  |  |  | 0 |
| G252 | Other specified forms of tremor |  |  |  | 0 |
| G254 | Drug-induced chorea |  |  |  | 0 |
| G255 | Other chorea |  |  |  | 0 |
| G258 | Other specified extrapyramidal and movement disorders |  |  |  | 0 |
| G259 | Extrapyramidal and movement disorder, unspecified |  |  |  | 0 |
| G26 | Extrapyramidal and movement disorders in diseases classified elsewhere |  |  |  | 0 |
| Y467 | Antiparkinsonism drugs |  |  |  | 0 |
| Y468 | Antispasticity drugs |  |  |  | 0 |
| G60 | Hereditary and idiopathic neuropathy |  |  |  | 0 |
| G600 | Hereditary motor and sensory neuropathy |  |  |  | 0 |
| G601 | Refsum disease |  |  |  | 0 |
| G602 | Neuropathy in association with hereditary ataxia |  |  |  | 0 |
| G603 | Idiopathic progressive neuropathy |  |  |  | 0 |
| G608 | Other hereditary and idiopathic neuropathies |  |  |  | 0 |
| G609 | Hereditary and idiopathic neuropathy, unspecified |  |  |  | 0 |
| G611 | Serum neuropathy |  |  |  | 0 |
| G618 | Other inflammatory polyneuropathies |  |  |  | 0 |
| G619 | Inflammatory polyneuropathy, unspecified |  |  |  | 0 |
| G62 | Other polyneuropathies |  |  |  | 0 |
| G620 | Drug-induced polyneuropathy |  |  |  | 0 |
| G621 | Alcoholic polyneuropathy |  |  |  | 0 |
| G622 | Polyneuropathy due to other toxic agents |  |  |  | 0 |
| G628 | Other specified polyneuropathies |  |  |  | 0 |
| G629 | Polyneuropathy, unspecified |  |  |  | 0 |
| G63 | Polyneuropathy in diseases classified elsewhere |  |  |  | 0 |
| G630 | Polyneuropathy in infectious and parasitic diseases classified elsewhere |  |  |  | 0 |
| G631 | Polyneuropathy in neoplastic disease |  |  |  | 0 |
| G632 | Diabetic polyneuropathy |  |  |  | 0 |
| G633 | Polyneuropathy in other endocrine and metabolic diseases |  |  |  | 0 |
| G634 | Polyneuropathy in nutritional deficiency |  |  |  | 0 |
| G635 | Polyneuropathy in systemic connective tissue disorders |  |  |  | 0 |
| G636 | Polyneuropathy in other musculoskeletal disorders |  |  |  | 0 |
| G638 | Polyneuropathy in other diseases classified elsewhere |  |  |  | 0 |
| G31 | Other degenerative diseases of nervous system, not elsewhere classified |  |  |  | 0 |
| G310 | Circumscribed brain atrophy |  |  |  | 0 |
| G311 | Senile degeneration of brain, not elsewhere classified |  |  |  | 0 |
| G312 | Degeneration of nervous system due to alcohol |  |  |  | 0 |
| G318 | Other specified degenerative diseases of nervous system |  |  |  | 0 |
| G319 | Degenerative disease of nervous system, unspecified |  |  |  | 0 |
| G32 | Other degenerative disorders of nervous system in diseases classified elsewhere |  |  |  | 0 |
| G320 | Subacute combined degeneration of spinal cord in diseases classified elsewhere |  |  |  | 0 |
| G328 | Other specified degenerative disorders of nervous system in diseases classified elsewhere |  |  |  | 0 |
| G834 | Cauda equina syndrome |  |  |  | 0 |
| G835 | Locked-in syndrome |  |  |  | 0 |
| G838 | Other specified paralytic syndromes |  |  |  | 0 |
| G839 | Paralytic syndrome, unspecified |  |  |  | 0 |
| G900 | Idiopathic peripheral autonomic neuropathy |  |  |  | 0 |
| G901 | Familial dysautonomia [Riley-Day] |  |  |  | 0 |
| G904 | Autonomic dysreflexia |  |  |  | 0 |
| G908 | Other disorders of autonomic nervous system |  |  |  | 0 |
| G909 | Disorder of autonomic nervous system, unspecified |  |  |  | 0 |
| G95 | Other diseases of spinal cord |  |  |  | 0 |
| G950 | Syringomyelia and syringobulbia |  |  |  | 0 |
| G951 | Vascular myelopathies |  |  |  | 0 |
| G951 | Vascular myelopathies |  |  |  | 0 |
| G952 | Cord compression, unspecified |  |  |  | 0 |
| G958 | Other specified diseases of spinal cord |  |  |  | 0 |
| G959 | Disease of spinal cord, unspecified |  |  |  | 0 |
| G99 | Other disorders of nervous system in diseases classified elsewhere |  |  |  | 0 |
| G990 | Autonomic neuropathy in endocrine and metabolic diseases |  |  |  | 0 |
| G991 | Other disorders of autonomic nervous system in other diseases classified elsewhere |  |  |  | 0 |
| G992 | Myelopathy in diseases classified elsewhere |  |  |  | 0 |
| G998 | Other specified disorders of nervous system in diseases classified elsewhere |  |  |  | 0 |
| **Perinatal conditions** | | | | | |
| P210 | Severe birth asphyxia | Severe birth asphyxia |  |  | 0 |
| P57 | Kernicterus | Perinatal Brain Damage |  |  | 0 |
| P570 | Kernicterus due to isoimmunization | Perinatal Brain Damage |  |  | 0 |
| P578 | Other specified kernicterus | Perinatal Brain Damage |  |  | 0 |
| P579 | Kernicterus, unspecified | Perinatal Brain Damage |  |  | 0 |
| P90 | Convulsions of newborn | Perinatal Brain Damage |  |  | 0 |
| P912 | Neonatal cerebral leukomalacia | Perinatal Brain Damage |  |  | 0 |
| G46 | Vascular syndromes of brain in cerebrovascular diseases | Perinatal Brain Damage | Perinatal stroke |  | 1 |
| G460 | Middle cerebral artery syndrome | Perinatal Brain Damage | Perinatal stroke |  | 1 |
| G461 | Anterior cerebral artery syndrome | Perinatal Brain Damage | Perinatal stroke |  | 1 |
| G462 | Posterior cerebral artery syndrome | Perinatal Brain Damage | Perinatal stroke |  | 1 |
| G463 | Brain stem stroke syndrome | Perinatal Brain Damage | Perinatal stroke |  | 1 |
| G464 | Cerebellar stroke syndrome | Perinatal Brain Damage | Perinatal stroke |  | 1 |
| G465 | Pure motor lacunar syndrome | Perinatal Brain Damage | Perinatal stroke |  | 1 |
| G466 | Pure sensory lacunar syndrome | Perinatal Brain Damage | Perinatal stroke |  | 1 |
| G467 | Other lacunar syndromes | Perinatal Brain Damage | Perinatal stroke |  | 1 |
| G468 | Other vascular syndromes of brain in cerebrovascular diseases | Perinatal Brain Damage | Perinatal stroke |  | 1 |
| I60 | Subarachnoid haemorrhage | Perinatal Brain Damage | Perinatal stroke |  | 1 |
| I600 | Subarachnoid haemorrhage from carotid siphon and bifurcation | Perinatal Brain Damage | Perinatal stroke |  | 1 |
| I601 | Subarachnoid haemorrhage from middle cerebral artery | Perinatal Brain Damage | Perinatal stroke |  | 1 |
| I602 | Subarachnoid haemorrhage from anterior communicating artery | Perinatal Brain Damage | Perinatal stroke |  | 1 |
| I603 | Subarachnoid haemorrhage from posterior communicating artery | Perinatal Brain Damage | Perinatal stroke |  | 1 |
| I604 | Subarachnoid haemorrhage from basilar artery | Perinatal Brain Damage | Perinatal stroke |  | 1 |
| I605 | Subarachnoid haemorrhage from vertebral artery | Perinatal Brain Damage | Perinatal stroke |  | 1 |
| I606 | Subarachnoid haemorrhage from other intracranial arteries | Perinatal Brain Damage | Perinatal stroke |  | 1 |
| I607 | Subarachnoid haemorrhage from intracranial artery, unspecified | Perinatal Brain Damage | Perinatal stroke |  | 1 |
| I608 | Other subarachnoid haemorrhage | Perinatal Brain Damage | Perinatal stroke |  | 1 |
| I609 | Subarachnoid haemorrhage, unspecified | Perinatal Brain Damage | Perinatal stroke |  | 1 |
| I61 | Intracerebral haemorrhage | Perinatal Brain Damage | Perinatal stroke |  | 1 |
| I610 | Intracerebral haemorrhage in hemisphere, subcortical | Perinatal Brain Damage | Perinatal stroke |  | 1 |
| I611 | Intracerebral haemorrhage in hemisphere, cortical | Perinatal Brain Damage | Perinatal stroke |  | 1 |
| I612 | Intracerebral haemorrhage in hemisphere, unspecified | Perinatal Brain Damage | Perinatal stroke |  | 1 |
| I613 | Intracerebral haemorrhage in brain stem | Perinatal Brain Damage | Perinatal stroke |  | 1 |
| I614 | Intracerebral haemorrhage in cerebellum | Perinatal Brain Damage | Perinatal stroke |  | 1 |
| I615 | Intracerebral haemorrhage, intraventricular | Perinatal Brain Damage | Perinatal stroke |  | 1 |
| I616 | Intracerebral haemorrhage, multiple localized | Perinatal Brain Damage | Perinatal stroke |  | 1 |
| I618 | Other intracerebral haemorrhage | Perinatal Brain Damage | Perinatal stroke |  | 1 |
| I619 | Intracerebral haemorrhage, unspecified | Perinatal Brain Damage | Perinatal stroke |  | 1 |
| I63 | Cerebral infarction | Perinatal Brain Damage | Perinatal stroke |  | 1 |
| I630 | Cerebral infarction due to thrombosis of precerebral arteries | Perinatal Brain Damage | Perinatal stroke |  | 1 |
| I631 | Cerebral infarction due to embolism of precerebral arteries | Perinatal Brain Damage | Perinatal stroke |  | 1 |
| I632 | Cerebral infarction due to unspecified occlusion or stenosis of precerebral arteries | Perinatal Brain Damage | Perinatal stroke |  | 1 |
| I633 | Cerebral infarction due to thrombosis of cerebral arteries | Perinatal Brain Damage | Perinatal stroke |  | 1 |
| I634 | Cerebral infarction due to embolism of cerebral arteries | Perinatal Brain Damage | Perinatal stroke |  | 1 |
| I635 | Cerebral infarction due to unspecified occlusion or stenosis of cerebral arteries | Perinatal Brain Damage | Perinatal stroke |  | 1 |
| I636 | Cerebral infarction due to cerebral venous thrombosis, nonpyogenic | Perinatal Brain Damage | Perinatal stroke |  | 1 |
| I638 | Other cerebral infarction | Perinatal Brain Damage | Perinatal stroke |  | 1 |
| I639 | Cerebral infarction, unspecified | Perinatal Brain Damage | Perinatal stroke |  | 1 |
| I64 | Stroke, not specified as haemorrhage or infarction | Perinatal Brain Damage | Perinatal stroke |  | 1 |
| I670 | Dissection of cerebral arteries, nonruptured | Perinatal Brain Damage | Perinatal stroke |  | 1 |
| I671 | Cerebral aneurysm, nonruptured | Perinatal Brain Damage | Perinatal stroke |  | 1 |
| I673 | Progressive vascular leukoencephalopathy | Perinatal Brain Damage | Perinatal stroke |  | 1 |
| I675 | Moyamoya disease | Perinatal Brain Damage | Perinatal stroke |  | 1 |
| I676 | Nonpyogenic thrombosis of intracranial venous system | Perinatal Brain Damage | Perinatal stroke |  | 1 |
| I677 | Cerebral arteritis, not elsewhere classified | Perinatal Brain Damage | Perinatal stroke |  | 1 |
| I678 | Other specified cerebrovascular diseases | Perinatal Brain Damage | Perinatal stroke |  | 1 |
| I679 | Cerebrovascular disease, unspecified | Perinatal Brain Damage | Perinatal stroke |  | 1 |
| I690 | Sequelae of subarachnoid haemorrhage | Perinatal Brain Damage | Perinatal stroke |  | 1 |
| I691 | Sequelae of intracerebral haemorrhage | Perinatal Brain Damage | Perinatal stroke |  | 1 |
| I693 | Sequelae of cerebral infarction | Perinatal Brain Damage | Perinatal stroke |  | 1 |
| I694 | Sequelae of stroke, not specified as haemorrhage or infarction | Perinatal Brain Damage | Perinatal stroke |  | 1 |
| P524 | Intracerebral (nontraumatic) haemorrhage of fetus and newborn | Perinatal Brain Damage | Perinatal stroke |  | 0 |
| P525 | Subarachnoid (nontraumatic) haemorrhage of fetus and newborn | Perinatal Brain Damage | Perinatal stroke |  | 0 |
| P526 | Cerebellar (nontraumatic) and posterior fossa haemorrhage of fetus and newborn | Perinatal Brain Damage | Perinatal stroke |  | 0 |
| P910 | Neonatal cerebral ischaemia | Perinatal Brain Damage | Perinatal stroke |  | 0 |
| P911 | Acquired periventricular cysts of newborn | Perinatal Brain Damage | Perinatal stroke |  | 0 |
| G91 | Hydrocephalus | Perinatal Brain Damage | Intracranial haemorrhage |  | 1 |
| G910 | Communicating hydrocephalus | Perinatal Brain Damage | Intracranial haemorrhage |  | 1 |
| G911 | Obstructive hydrocephalus | Perinatal Brain Damage | Intracranial haemorrhage |  | 1 |
| G912 | Normal-pressure hydrocephalus | Perinatal Brain Damage | Intracranial haemorrhage |  | 1 |
| G913 | Post-traumatic hydrocephalus, unspecified | Perinatal Brain Damage | Intracranial haemorrhage |  | 1 |
| G918 | Other hydrocephalus | Perinatal Brain Damage | Intracranial haemorrhage |  | 1 |
| G919 | Hydrocephalus, unspecified | Perinatal Brain Damage | Intracranial haemorrhage |  | 1 |
| G940 | Hydrocephalus in infectious and parasitic diseases classified elsewhere | Perinatal Brain Damage | Intracranial haemorrhage |  | 1 |
| G941 | Hydrocephalus in neoplastic disease | Perinatal Brain Damage | Intracranial haemorrhage |  | 1 |
| G942 | Hydrocephalus in other diseases classified elsewhere | Perinatal Brain Damage | Intracranial haemorrhage |  | 1 |
| G972 | Intracranial hypotension following ventricular shunting | Perinatal Brain Damage | Intracranial haemorrhage |  | 1 |
| P100 | Subdural haemorrhage due to birth injury | Perinatal Brain Damage | Intracranial haemorrhage |  | 0 |
| P101 | Cerebral haemorrhage due to birth injury | Perinatal Brain Damage | Intracranial haemorrhage |  | 0 |
| P102 | Intraventricular haemorrhage due to birth injury | Perinatal Brain Damage | Intracranial haemorrhage |  | 0 |
| P103 | Subarachnoid haemorrhage due to birth injury | Perinatal Brain Damage | Intracranial haemorrhage |  | 0 |
| P104 | Tentorial tear due to birth injury | Perinatal Brain Damage | Intracranial haemorrhage |  | 0 |
| P108 | Other intracranial lacerations and haemorrhages due to birth injury | Perinatal Brain Damage | Intracranial haemorrhage |  | 0 |
| P109 | Unspecified intracranial laceration and haemorrhage due to birth injury | Perinatal Brain Damage | Intracranial haemorrhage |  | 0 |
| P522 | Intraventricular (nontraumatic) haemorrhage, grade 3 and grade 4, of fetus and newborn | Perinatal Brain Damage | Intracranial haemorrhage |  | 0 |
| P523 | Unspecified intraventricular (nontraumatic) haemorrhage of fetus and newborn | Perinatal Brain Damage | Intracranial haemorrhage |  | 0 |
| P528 | Other intracranial (nontraumatic) haemorrhages of fetus and newborn | Perinatal Brain Damage | Intracranial haemorrhage |  | 0 |
| P529 | Intracranial (nontraumatic) haemorrhage of fetus and newborn, unspecified | Perinatal Brain Damage | Intracranial haemorrhage |  | 0 |
| P917 | Acquired hydrocephalus of newborn | Perinatal Brain Damage | Intracranial haemorrhage |  | 0 |
| T850 | Mechanical complication of ventricular intracranial (communicating) shunt | Perinatal Brain Damage | Intracranial haemorrhage |  | 1 |
| Z982 | Presence of cerebrospinal fluid drainage device | Perinatal Brain Damage | Intracranial haemorrhage |  | 1 |
| P915 | Neonatal coma | Perinatal Brain Damage |  |  | 0 |
| P916 | Hypoxic ischaemic encephalopathy of newborn | Perinatal Brain Damage |  |  | 0 |
| A170 | Tuberculous meningitis | Perinatal Brain Damage | CNS infection |  | 1 |
| A171 | Meningeal tuberculoma | Perinatal Brain Damage | CNS infection |  | 1 |
| A203 | Plague meningitis | Perinatal Brain Damage | CNS infection |  | 1 |
| A321 | Listerial meningitis and meningoencephalitis | Perinatal Brain Damage | CNS infection |  | 1 |
| A390 | Meningococcal meningitis | Perinatal Brain Damage | CNS infection |  | 1 |
| A872 | Lymphocytic choriomeningitis | Perinatal Brain Damage | CNS infection |  | 1 |
| B003 | Herpesviral meningitis | Perinatal Brain Damage | CNS infection |  | 1 |
| B004 | Herpesviral encephalitis | Perinatal Brain Damage | CNS infection |  | 1 |
| B007 | Disseminated herpesviral disease | Perinatal Brain Damage | CNS infection |  | 1 |
| B010 | Varicella meningitis | Perinatal Brain Damage | CNS infection |  | 1 |
| B011 | Varicella encephalitis | Perinatal Brain Damage | CNS infection |  | 1 |
| B018 | Varicella with other complications | Perinatal Brain Damage | CNS infection |  | 1 |
| B375 | Candidal meningitis | Perinatal Brain Damage | CNS infection |  | 1 |
| G00 | Bacterial meningitis, not elsewhere classified | Perinatal Brain Damage | CNS infection |  | 1 |
| G000 | Haemophilus meningitis | Perinatal Brain Damage | CNS infection |  | 1 |
| G001 | Pneumococcal meningitis | Perinatal Brain Damage | CNS infection |  | 1 |
| G002 | Streptococcal meningitis | Perinatal Brain Damage | CNS infection |  | 1 |
| G003 | Staphylococcal meningitis | Perinatal Brain Damage | CNS infection |  | 1 |
| G008 | Other bacterial meningitis | Perinatal Brain Damage | CNS infection |  | 1 |
| G009 | Bacterial meningitis, unspecified | Perinatal Brain Damage | CNS infection |  | 1 |
| G01 | Meningitis in bacterial diseases classified elsewhere | Perinatal Brain Damage | CNS infection |  | 1 |
| G021 | Meningitis in mycoses | Perinatal Brain Damage | CNS infection |  | 1 |
| G028 | Meningitis in other specified infectious and parasitic diseases classified elsewhere | Perinatal Brain Damage | CNS infection |  | 1 |
| G03 | Meningitis due to other and unspecified causes | Perinatal Brain Damage | CNS infection |  | 1 |
| G030 | Nonpyogenic meningitis | Perinatal Brain Damage | CNS infection |  | 1 |
| G031 | Chronic meningitis | Perinatal Brain Damage | CNS infection |  | 1 |
| G032 | Benign recurrent meningitis [Mollaret] | Perinatal Brain Damage | CNS infection |  | 1 |
| G038 | Meningitis due to other specified causes | Perinatal Brain Damage | CNS infection |  | 1 |
| G039 | Meningitis, unspecified | Perinatal Brain Damage | CNS infection |  | 1 |
| G05 | Encephalitis, myelitis and encephalomyelitis in diseases classified elsewhere | Perinatal Brain Damage | CNS infection |  | 1 |
| G050 | Encephalitis, myelitis and encephalomyelitis in bacterial diseases classified elsewhere | Perinatal Brain Damage | CNS infection |  | 1 |
| G051 | Encephalitis, myelitis and encephalomyelitis in viral diseases classified elsewhere | Perinatal Brain Damage | CNS infection |  | 1 |
| G052 | Encephalitis, myelitis and encephalomyelitis in other infectious and parasitic diseases classified elsewhere | Perinatal Brain Damage | CNS infection |  | 1 |
| G058 | Encephalitis, myelitis and encephalomyelitis in other diseases classified elsewhere | Perinatal Brain Damage | CNS infection |  | 1 |
| P352 | Congenital herpesviral [herpes simplex] infection | Perinatal Brain Damage | CNS infection |  | 0 |
| P044 | Fetus and newborn affected by maternal use of drugs of addiction |  |  |  | 0 |
| P961 | Neonatal withdrawal symptoms from maternal use of drugs of addiction |  |  |  | 0 |
| A50 | Congenital syphilis |  |  |  | 0 |
| A500 | Early congenital syphilis, symptomatic |  |  |  | 0 |
| A501 | Early congenital syphilis, latent |  |  |  | 0 |
| A502 | Early congenital syphilis, unspecified |  |  |  | 0 |
| A503 | Late congenital syphilitic oculopathy |  |  |  | 0 |
| A504 | Late congenital neurosyphilis [juvenile neurosyphilis] |  |  |  | 0 |
| A505 | Other late congenital syphilis, symptomatic |  |  |  | 0 |
| A506 | Late congenital syphilis, latent |  |  |  | 0 |
| A507 | Late congenital syphilis, unspecified |  |  |  | 0 |
| A509 | Congenital syphilis, unspecified |  |  |  | 0 |
| B060 | Rubella with neurological complications |  |  |  | 1 |
| B069 | Rubella without complication |  |  |  | 1 |
| B582 | Toxoplasma meningoencephalitis |  |  |  | 1 |
| P350 | Congenital rubella syndrome |  |  |  | 0 |
| P351 | Congenital cytomegalovirus infection |  |  |  | 0 |
| P358 | Other congenital viral diseases |  |  |  | 0 |
| P359 | Congenital viral disease, unspecified |  |  |  | 0 |
| P370 | Congenital tuberculosis |  |  |  | 0 |
| P371 | Congenital toxoplasmosis |  |  |  | 0 |

^†^ All codes contribute to classification of any neurodisability; ^‡^ Use of code is restricted to admissions occurring within the first 28 days of life.

ADHD: Attention Deficit Hyperactivity Disorder; CNS: Central Nervous System; ICD-10: International Classification of Diseases 10^th^ revision.

**Appendix Table 2. Special Educational Needs and Disability (SEND) types of need used to identify intellectual disability and autism in school records.**

| **Neurodisability** | **SEND type of need** | **Description** |
| --- | --- | --- |
| Intellectual disability  Note: Intellectual disability is commonly referred to as learning disability in England. | PMLD | **Profound & Multiple Learning Difficulty**  Where children are likely to have severe and complex learning difficulties as well as a physical disability or sensory impairment.^3^ |
|  | SLD | **Severe Learning Difficulty**  Where children are likely to need support in all areas of the curriculum and associated difficulties with mobility and communication.^3^ |
|  | Exclusions: MLD | **Moderate Learning Difficulty**  Excluded due to its status as a highly heterogeneous educational label that lacks clear diagnostic thresholds and frequently overlaps with general low academic attainment.^4^ |
| Autism | ASD | **Autistic Spectrum Disorder** Where children are likely to have particular difficulties with social interaction. They may also experience difficulties with language, communication and imagination, which can impact on how they relate to others.^3^ |
