## supporting_information for "Mortality risk between ages 11 and 22 years among young people with neurodisability in England: a national cohort study using linked health and education data"

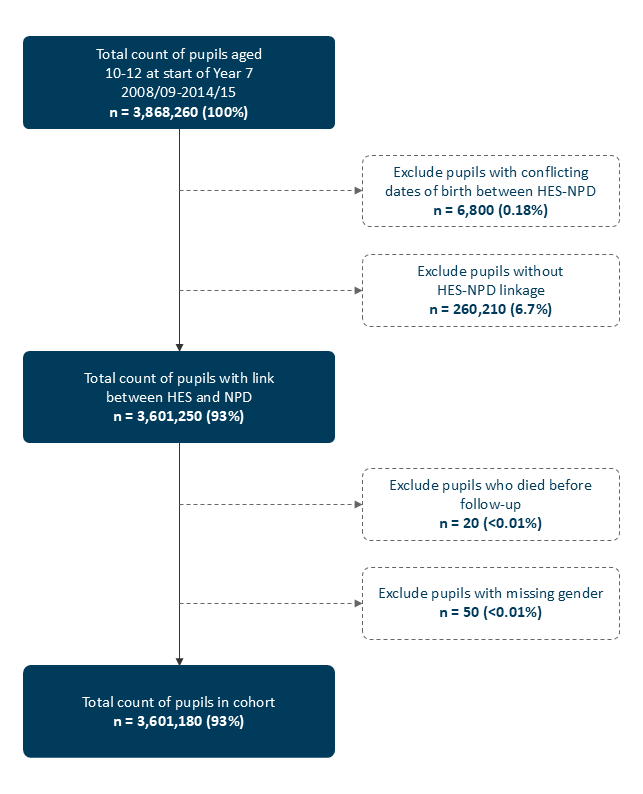

**Figure S1. Flow diagram of study cohort derivation.** Counts are rounded to the nearest 10 to protect confidentiality and percentages are calculated from rounded counts. Totals may not sum exactly due to rounding. Exact counts for the initial and final cohort are reported in the main text.

HES: Hospital Episode Statistics; NPD: National Pupil Database

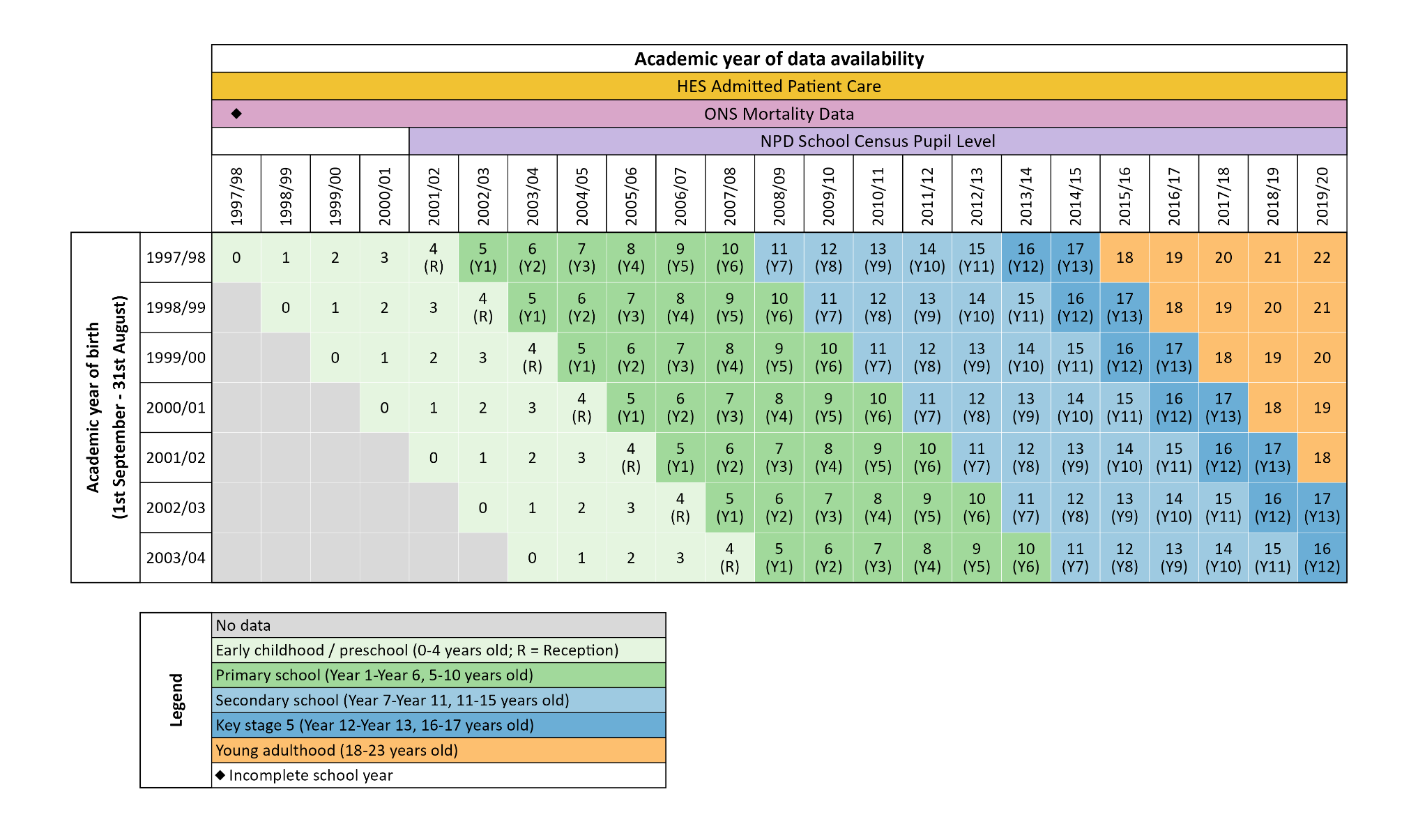

**Figure S2. Overview of the study cohorts, expected age at the start of each academic year, and availability of linked data sources.** Cells show the expected age at the start of the academic year and the corresponding school year. Study follow-up begins at entry to secondary school (Year 7, age 11 years). Colours indicate educational stage, as defined in the legend

A&E: Accident and Emergency; HES: Hospital Episode Statistics; NPD: National Pupil Database; ONS: Office for National Statistics.

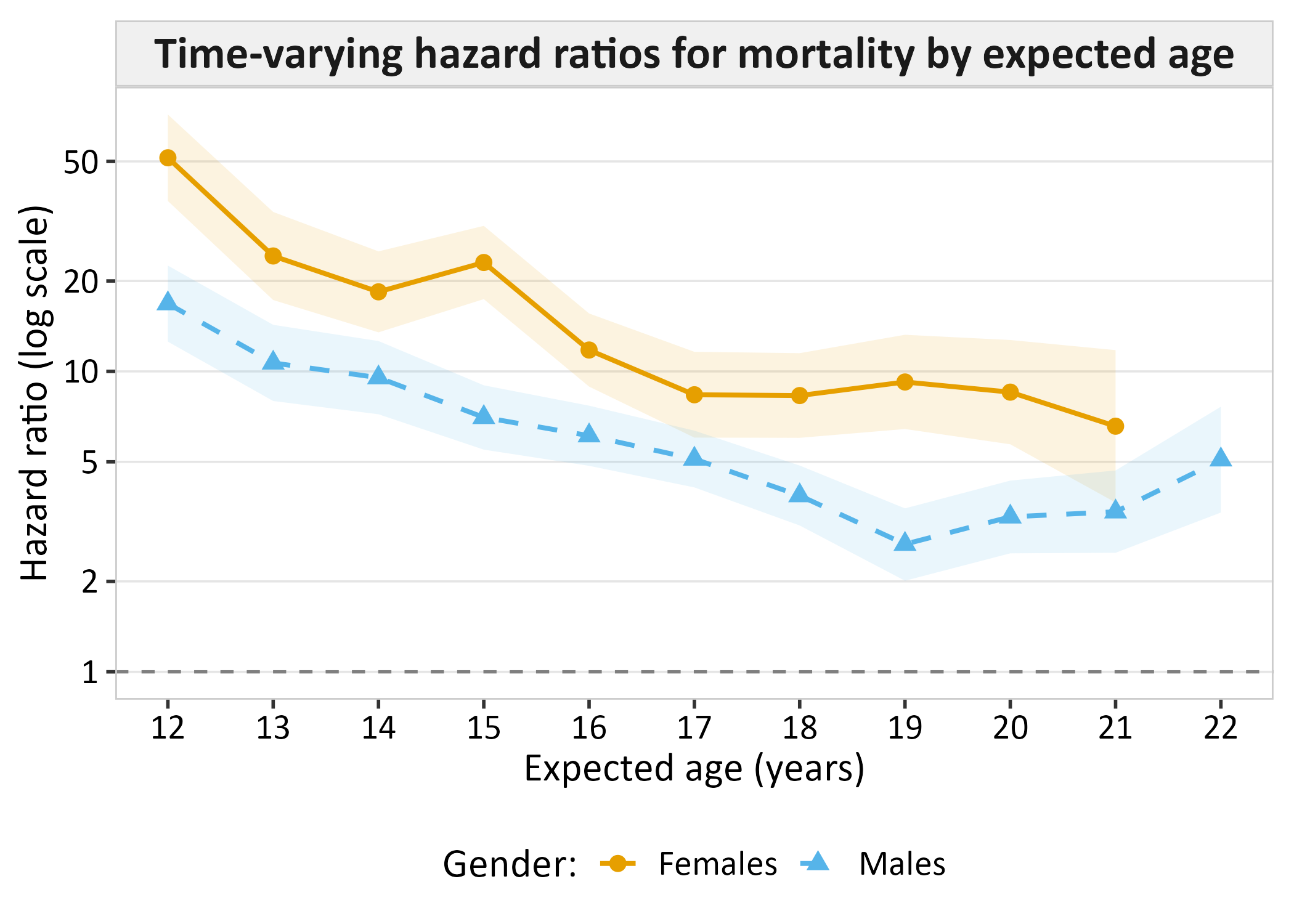

**Figure S3.** **Time-varying hazard ratios for mortality by expected age.** Hazard ratios were estimated using Cox proportional hazards models with interaction terms between exposure and time intervals to account for non-proportional hazards. Expected age (years) represents approximate age by school year from the start of Year 7 follow-up (pupils aged 10-12 years at baseline) and estimates reflect risk within the preceding year. Estimates for females at age 22 are not reported due to small counts.

**Table S1. Characteristics of young people not linked to Hospital Episode Statistics (HES) records.**

| **Characteristic**^‡^ | **Unlinked pupils**  **260,210 (100%)^a^** | **Linked pupils**  **3,601,180 (100%)^a^** |
| --- | --- | --- |
| **Gender** |  |  |
| Females | 134,436 (52%) | 1,747,730 (49%) |
| Males | 125,774 (48%) | 1,853,450 (51%) |
| **Ethnic group** |  |  |
| White | 183,757 (71%) | 2,863,263 (80%) |
| Asian | 33,374 (13%) | 312,257 (8.7%) |
| Black | 19,121 (7.3%) | 167,526 (4.7%) |
| Chinese | 1,664 (0.64%) | 11,305 (0.31%) |
| Mixed | 11,917 (4.6%) | 150,906 (4.2%) |
| Any other ethnic group | 6,186 (2.4%) | 44,490 (1.2%) |
| Missing | 4,191 (1.6%) | 51,433 (1.4%) |
| **First language** |  |  |
| English | 198,618 (76%) | 3,128,212 (87%) |
| Other | 59,814 (23%) | 452,810 (13%) |
| Missing | 1,778 (0.68%) | 20,158 (0.56%) |
| **Recorded as FSM eligible** |  |  |
| No | 215,940 (83%)^†^ | 2,968,540 (82%)^†^ |
| Yes | 44,280 (17%)^†^ | 632,630 (18%)^†^ |
| Missing | ^*^ | ^*^ |
| **IDACI quintile** |  |  |
| Q1 (most deprived) | 68,667 (26%) | 864,450 (24%) |
| Q2 | 52,051 (20%) | 727,270 (20%) |
| Q3 | 45,998 (18%) | 676,030 (19%) |
| Q4 | 45,821 (18%) | 667,397 (19%) |
| Q5 (least deprived) | 46,076 (18%) | 648,763 (18%) |
| Missing | 1,597 (0.61%) | 17,270 (0.48%) |
| **Year 7 school** |  |  |
| Mainstream | 256,879 (99%) | 3,537,481 (98%) |
| Special | 3,331 (1.3%) | 63,699 (1.8%) |
| **Ever any SEND provision** |  |  |
| No | 175,696 (68%) | 2,288,591 (64%) |
| Yes | 84,514 (32%) | 1,312,589 (36%) |
| **Highest SEND provision** |  |  |
| SEN support | 77,423 (30%) | 1,183,684 (33%) |
| EHCP | 7,091 (2.7%) | 128,905 (3.6%) |

^a^ n (% of all); ^*^ Counts are suppressed where n <10 to protect confidentiality; ^†^ Counts are rounded to the nearest 10 to protect confidentiality and percentages are calculated from rounded counts, totals may not sum exactly due to rounding; ^‡^ Characteristics are presented for pupils with recorded gender.

EHCP: Education, Health and Care plan; FSM: Free school meals; IDACI: Income Deprivation Affecting Children Index; SEN: Special Educational Needs; SEND: Special Educational Needs and Disabilities.

**Table S2. Distribution of underlying causes of death by ICD-10 chapter among deaths due to medical causes.**

|  | **Girls** | |  | **Boys** | |
| --- | --- | --- | --- | --- | --- |
| **ICD-10 chapter:  Underlying cause of death** | **No  neurodisability  n (%)^a^** | **Any  neurodisability  n (%)^a^** |  | **No  neurodisability  n (%)^a^** | **Any  neurodisability  n (%)^a^** |
| Total deaths from medical causes | 714 | 466 |  | 854 | 681 |
| Infectious and parasitic diseases | 36 (5.0%) | 18 (3.9%) |  | 30 (3.5%) | 20 (2.9%) |
| Neoplasms | 284 (40%) | 47 (10%) |  | 338 (40%) | 86 (13%) |
| Blood and immune disorders | 16 (2.2%) | 14 (3.0%) |  | 28 (3.3%) | 11 (1.6%) |
| Endocrine and metabolic | 68 (10%) | 44 (9.4%) |  | 44 (5.2%) | 67 (10%) |
| Mental and behavioural | ^*^ | 13 (2.8%) |  | ^*^ | ^*^ |
| Nervous system | 66 (9.2%) | 125 (27%) |  | 83 (10%) | 237 (35%) |
| Circulatory system | 102 (14%) | 16 (3.4%) |  | 158 (19%) | 40 (5.9%) |
| Respiratory system | 60 (8.4%) | 57 (12%) |  | 56 (6.6%) | 78 (11%) |
| Digestive system | 13 (1.8%) | 24 (5.2%) |  | 17 (2.0%) | 32 (4.7%) |
| Musculoskeletal | ^*^ | 13 (2.8%) |  | ^*^ | ^*^ |
| Genitourinary | ^*^ | 12 (2.6%) |  | ^*^ | ^*^ |
| Congenital malformations | 25 (3.5%) | 66 (14%) |  | 42 (4.9%) | 75 (11%) |
| Symptoms & abnormal findings | 23 (3.2%) | ^*^ |  | 33 (3.9%) | ^*^ |
| External causes^‡^ | ^*^ | ^*^ |  | 12 (1.4%) | ^*^ |
| Other ICD-10 chapters^§^ | ^*^ | ^*^ |  | ^*^ | ^*^ |

^a^ n (% of all medical deaths); ^*^ Counts are suppressed where n < 10 to protect confidentiality; ^‡^ Some medical causes of death were coded to ICD-10 Chapter XX (“External causes”), shown here for transparency; ^§^ Includes eye/ear, skin, pregnancy and childbirth, perinatal conditions, and other ICD-10 chapters with very few deaths.

ICD-10: International Classification of Diseases 10^th^ revision.

**Table S3. Prevalence before age 11 years and cumulative mortality risk between ages 11 and 22 years by neurodisability subgroup.**

|  | **Girls** | | | |  | **Boys** | | | |
| --- | --- | --- | --- | --- | --- | --- | --- | --- | --- |
| **Neurodisability subgroup^‡^** | **N** | **Prevalence before  age 11 years** | **Deaths** | **Cumulative risk  (95% CI)** |  | **N** | **Prevalence before  age 11 years** | **Deaths** | **Cumulative risk  (95% CI)** |
| **No neurodisability** | 1,702,771 | 97.4% | 1471 | 0.14% (0.13-0.15%) |  | 1,754,545 | 94.7% | 2779 | 0.28% (0.27-0.29%) |
| **Any neurodisability** | 44,959 | 2.6% | 514 | 1.6% (1.4-1.8%) |  | 98,905 | 5.3% | 801 | 1.3% (1.2-1.4%) |
| **Neurodevelopmental conditions** |  |  |  |  |  |  |  |  |  |
| Intellectual disability (HES and NPD) | 17,143 | 0.98% | 397 | 3.1% (2.7-3.5%) |  | 29,680 | 1.6% | 533 | 2.6% (2.3-2.9%) |
| Developmental Delay | 6,664 | 0.38% | 237 | 5.0% (4.2-5.9%) |  | 12,196 | 0.66% | 285 | 3.5% (3.0-4.0%) |
| Autism (HES and NPD) | 7,744 | 0.44% | 24 | 0.52% (0.23-0.82%) |  | 43,307 | 2.3% | 106 | 0.38% (0.29-0.46%) |
| Attention Deficit Hyperactivity Disorder | 1,045 | 0.06% | ^*^ | ^*^ |  | 5,480 | 0.30% | 29 | 1.0% (0.51-1.5%) |
| **Complex neurological conditions** |  |  |  |  |  |  |  |  |  |
| Cerebral Palsy | 4,549 | 0.26% | 230 | 6.5% (5.5-7.5%) |  | 6,136 | 0.33% | 265 | 6.3% (5.4-7.3%) |
| Epilepsy | 8,400 | 0.48% | 270 | 4.2% (3.6-4.9%) |  | 10,870 | 0.59% | 325 | 4.3% (3.8-4.9%) |
| **Congenital conditions** |  |  |  |  |  |  |  |  |  |
| Any congenital condition | 9,133 | 0.52% | 262 | 4.1% (3.5-4.8%) |  | 11,707 | 0.63% | 328 | 4.3% (3.7-4.8%) |
| Inherited metabolic conditions | 860 | 0.05% | 57 | 9.1% (6.2-12.0%) |  | 1,244 | 0.07% | 90 | 10.6% (7.6-13.6%) |
| Any chromosomal condition | 2,100 | 0.12% | 51 | 3.2% (2.0-4.4%) |  | 2,386 | 0.13% | 48 | 3.0% (2.1-4.0%) |
| Down syndrome | 1,504 | 0.09% | 12 | 1.4% (0.08-2.8%) |  | 1,767 | 0.10% | 17 | 1.7% (0.75-2.6%) |
| Anomalies of sex chromosome | 616 | 0.04% | 27 | 5.4% (3.3-7.4%) |  | 959 | 0.05% | 26 | 3.5% (2.0-4.9%) |
| Anomalies of the CNS | 3,484 | 0.20% | 143 | 6.2% (4.8-7.7%) |  | 4,186 | 0.23% | 167 | 6.2% (5.0-7.4%) |
| Microcephaly | 1,386 | 0.08% | 86 | 9.1% (6.5-11.7%) |  | 1,409 | 0.08% | 103 | 10.2% (8.0-12.4%) |
| Congenital hydrocephalus | 605 | 0.03% | 26 | 5.9% (2.8-8.9%) |  | 918 | 0.05% | 30 | 5.8% (3.2-8.4%) |
| Spina bifida | 411 | 0.02% | 13 | 4.3% (1.8-6.8%) |  | 424 | 0.02% | ^*^ | ^*^ |
| **High-risk conditions affecting the brain** |  |  |  |  |  |  |  |  |  |
| Hydrocephalus | 1,355 | 0.08% | 65 | 6.3% (4.5-8.1%) |  | 1,975 | 0.11% | 71 | 5.4% (3.8-6.9%) |
| Paediatric Stroke | 662 | 0.04% | 24 | 4.6% (2.7-6.5%) |  | 851 | 0.05% | 28 | 5.7% (2.8-8.4%) |
| CNS tumours | 609 | 0.03% | 37 | 7.3% (4.8-9.8%) |  | 705 | 0.04% | 42 | 7.4% (5.0-9.8%) |
| Inflammatory conditions of the brain | 3,434 | 0.20% | 22 | 0.72% (0.40-1.0%) |  | 4,835 | 0.26% | 40 | 1.2% (0.75-1.6%) |
| Meningitis | 2,430 | 0.14% | 10 | 0.52% (0.18-0.86%) |  | 3,588 | 0.19% | 20 | 0.86% (0.42-1.3%) |
| **Sensory impairments** |  |  |  |  |  |  |  |  |  |
| Hearing impairment | 2,287 | 0.13% | 29 | 2.1% (1.0-3.2%) |  | 2,822 | 0.15% | 42 | 2.5% (1.5-3.5%) |
| Visual impairment | 1,663 | 0.10% | 80 | 6.9% (4.6-9.2%) |  | 1,942 | 0.10% | 99 | 7.1% (5.4-8.9%) |
| **Impairment of motor function** | 2,084 | 0.12% | 117 | 7.9% (6.1-9.6%) |  | 2,758 | 0.15% | 208 | 11.6% (9.6-13.6%) |
| **Perinatal conditions** |  |  |  |  |  |  |  |  |  |
| Severe birth asphyxia | 1,503 | 0.09% | ^*^ | ^*^ |  | 2,291 | 0.12% | 11 | 0.67% (0.26-1.1%) |
| Perinatal Brain Damage | 6,367 | 0.36% | 114 | 2.3% (1.8-2.9%) |  | 9,041 | 0.49% | 133 | 2.5% (1.9-3.1%) |
| Perinatal stroke | 955 | 0.05% | 29 | 3.7% (2.3-5.2%) |  | 1,214 | 0.07% | 31 | 4.6% (2.1-7.0%) |
| Intracranial haemorrhage | 1,873 | 0.11% | 70 | 5.1% (3.6-6.5%) |  | 2,773 | 0.15% | 76 | 4.4% (3.1-5.8%) |
| CNS infections (perinatal) | 2,624 | 0.15% | 13 | 0.60% (0.25-0.94%) |  | 3,844 | 0.21% | 21 | 0.83% (0.41-1.3%) |

^*^ Counts are suppressed where n < 10 to protect confidentiality and corresponding estimates are not reported; **^‡^** Neurodisability subgroups are non-mutually exclusive. Intellectual disability refers to cases defined using the primary case definition, including explicit diagnoses, high-risk conditions, and associated conditions. All subgroups are identified from HES data unless otherwise stated.

CI: Confidence Interval; CNS: Central Nervous System; HES: Hospital Episode Statistics; NPD: National Pupil Database.

**Table S4. Neurodisability subgroup prevalence before age 11 years in the study cohort compared with published population-based estimates.**

| **Neurodisability subgroup^†^** | **Cohort prevalence before age 11 years^‡^** | **Published population-based estimates^§^** |
| --- | --- | --- |
| Neurodisability | 4.0% | - 3.6% of children aged <11 years had hospital‐recorded neurodisability in England between 2003-2019.^1^ - 6.3% of children aged 5-9 years had a neurodevelopmental condition, based on Global Burden of Disease data 2019.^2^ |
| **Neurodevelopmental conditions** | | |
| Intellectual disability (with or without autism).   Note: Intellectual disability is commonly referred to as learning disability in England. | 1.30%  (HES or NPD) | - **2.5%** of children aged 5-16 years had a learning disability flag in their electronic medical records (based on clinical review of multi-agency Education, Health and Care assessments) at a large regional hospital in North East England between 2017-2019.^3^ - **0.14%** of children had Severe Learning Difficulty or Profound & Multiple Learning Difficulty recorded as a type of SEND (rising to 3.5% including Moderate Learning Difficulty) in state funded primary schools in England in 2015/16.^4^ - **1.3 - 3.8%** of children aged <16 years, ranging from the more explicitly defined 2007 NISALD survey data (1.3%) to the more crudely classified 2011 Census data (3.8%) in Northern Ireland.^5^ |
| Developmental delay | 0.52% |  |
| Autism (with or without intellectual disability) | 1.4%  (HES or NPD) | - **2.9%** of children aged 10-14 years had a diagnosis recorded in primary care data in England as of 2018.^6^ - **0.84%** of children had autism recorded as a type of SEND in state funded primary schools in England in the 2015/16 academic year.^4^ - **1.5%** of children aged 5-10 years, based on a survey of the mental health of children and young people in England in 2017.^7^ |
| Attention Deficit Hyperactivity Disorder (ADHD) | 0.18% | - **2.6%** of boys and **0.8%** of girls aged 5-10 years had a hyperactivity disorder, based on a survey of the mental health of children and young people in England in 2017.^7^ - **1.8%** (175 per 10,000) male and **0.38%** (37.7 per 10,000) female period prevalence in children aged 3-17 years with a diagnosis recorded in primary care records in the UK between 2000-2018.^8^ - **0.93%** of pharmacologically treated primary and secondary schoolchildren, based on linked education and health records in Scotland and Wales between 2009-2016.^9^ |
| **Complex neurologic conditions** | | |
| Cerebral palsy | 0.30% | - **0.22%** (22 per 10,000) sex-standardised period prevalence among children aged <10 years, using linked primary care and secondary care records in England between 2010-2015.^10^ - **0.34%** (34 per 10,000) period prevalence among children and young people aged <25 years, using linked primary care and hospital records in England between 2004-2014.^11^ - **0.27%** (27 per 10,000) prevalence among young people aged <25 years, identified through linkage between The Northern Ireland Cerebral Palsy Register and hospital data in Northern Ireland between 2004-2014.^12^ |
| Epilepsy | 0.54% | - **0.69%** prevalence among treated schoolchildren aged 4-18 years, using linked hospital records and primary care prescription data in Scotland between 2009-2013.^13^ - **0.61%** (61 per 10,000) sex-standardised period prevalence among children aged <10 years, using linked primary care and secondary care records in England between 2010-2015.^10^ - **0.30% - 0.33%** (30.1/10,000 in CPRD Gold and 32.9/10,000 in CPRD Aurum) prevalence among children aged 5-9 years, using primary care records in England between 2013-2018.^14^ |
| **Congenital anomalies** | | |
| Inherited metabolic conditions | 0.06% | - **0.13%** (12.8 per 10,000) live birth prevalence in the West Midlands, England, using data from a population-based study of inherited metabolic disorders between 1999-2003.^15^ |
| Down syndrome | 0.09% | - **0.12%** (12.3 per 10,000) live birth prevalence, using linked data from NDSCR and HES in England between 1998–2013.^16^ - **0.12%** (11.6 per 10,000) live birth prevalence registered with NCARDRS in England in 2018.^17^ - **0.10%** (9.8 per 10,000) live birth prevalence in European countries participating in EUROCAT between 1995-2004.^18^ |
| Anomalies of sex chromosome | 0.04% | - **0.002%** (0.22 per 10,000) live birth prevalence of Klinefelter’s syndrome registered with NCARDRS in England in 2018.^17^ - **0.01%** (1.17 per 10,000) point prevalence of fragile X syndrome among children aged 5-10 years using primary care records in England in 2019.^19^ |
| Any central nervous system anomalies | 0.21% | - **0.09%** (9.2 per 10,000) live birth prevalence registered with NCARDRS in England in 2018.^17^ - **0.13%** (12.8 per 10,000) live birth prevalence in European countries participating in EUROCAT between 1995-2004.^18^ |
| Microcephaly | 0.08% | - **0.01%** (0.81 per 10,000) live birth prevalence registered with NCARDRS in England in 2018.^17^ - **0.02%** (2.3 per 10,000) live birth prevalence in European countries participating in EUROCAT between 1995-2004.^18^ |
| Congenital hydrocephalus | 0.04% | - **0.02%** (2.3 per 10,000) live birth prevalence registered with NCARDRS in England in 2018.^17^ - **0.03%** (3.2 per 10,000) live birth prevalence in European countries participating in EUROCAT between 1995-2004.^18^ |
| Spina bifida | 0.02% | - **0.02%** (2.2 per 10,000) live birth prevalence registered with NCARDRS in England in 2018.^17^ - **0.03%** (2.9 per 10,000) live birth prevalence in European countries participating in EUROCAT between 1995-2004.^18^ |
| **High-risk conditions affecting the brain** | | |
| Hydrocephalus | 0.09% | - **0.09%** (8.7 per 10,000) pooled prevalence among children aged <18 years in Europe, based on a systematic review and meta-analysis of international studies up to 2018.^20^ |
| Paediatric stroke | 0.04% | - **0.03%** (3 per 10,000) sex-standardised period prevalence of ischaemic stroke and intracerebral haemorrhage, respectively, among children aged <10 years, using linked primary care and secondary care records in England between 2010-2015.^10^ - **0.06%** (6.0 per 10,000) age-standardised prevalence among children aged <19 years in the UK in 2019.^21^ |
| Central nervous system tumours | 0.04% | - **0.03%** (3 per 10,000) sex-standardised period prevalence of primary malignancy in the brain among children aged <10 years, using linked primary care and secondary care records in England between 2010-2015.^10^ - **0.03%** (3.2 per 10,000) period prevalence among school children with a previous diagnosis of malignant neoplasms of the eye, brain, or other parts of the central nervous system, using linked education and health records in Scotland between 2009-2013.^22^ |
| Meningitis | 0.17% | - **0.15%** (15 per 10,000) sex-standardised period prevalence among children aged <10 years, using linked primary care and secondary care records in England between 2010-2015.^10^ |
| **Sensory impairment** | | |
| Hearing Impairment | 0.14% | - **0.09-0.17%** (9.1-16.5 per 10,000), rising to **0.11-0.21%** (10.7-20.5 per 10,000) when adjusted for under ascertainment, of permanent bilateral hearing impairment (≥40 decibel) among children aged 3-16 years, estimated from health and education sector sources in England between 1980-1995.^23^ - **3.01%** (301 per 10,000) sex-standardised period prevalence of deafness (all types and degrees, including mild and conductive hearing loss) among children aged <10 years, using linked primary care and secondary care records in England between 2010-2015.^10^ |
| Visual Impairment | 0.10% | - **0.09%** (8.89 per 10,000) cumulative incidence of any visual impairment among children up to 10, from a population-based register of visual impairment in southern England between 1984-1998.^24^ - **0.18%** (18 per 10,000) sex-standardised period prevalence of blindness (all causes and severities) among children aged <10 years, using linked primary care and secondary care records in England between 2010-2015.^10^ |
| **Impairment of motor function** | | |
| Impairment of motor function | 0.13% | - **0.06%** (5.8 per 10,000) period prevalence of neuromuscular diseases among children aged 5-9 years, based on primary care records in the UK in 2019.^25^ - **0.04%** (3.7 per 10,000) period prevalence of neuromuscular diseases among children aged <16 years in Yorkshire, England, based on clinically confirmed cases from regional specialist neurology and therapy databases in 2010.^26^ - **0.006%** (0.62 per 10,000) birth prevalence and **0.0002%** (0.02 per 10,000) population prevalence of Spinal Muscular Atrophy Type 1 (SMA1), based on HES data in England between 2008-2016.^27^ |
| **Perinatal conditions** | | |
| Severe birth asphyxia | 0.11% | - **0.24%** (24.0 per 10,000) live birth incidence of Hypoxic-ischaemic encephalopathy in England, using data from the NNRD in 2012.^28^ |
| Perinatal brain damage | 0.43% | - **0.48%** (48.4 per 10,000) live birth incidence, using data from the NNRD in 2012.^28^ |
| Perinatal stroke | 0.06% | - **0.01%** (1.1 per 10,000) live birth incidence in England, using data from the NNRD in 2012.^28^ |
| Intracranial haemorrhage | 0.13% | - **0.11%** (11.0 per 10,000) live birth incidence in England, using data from the NNRD in 2012.^28^ |
| Central nervous system infections | 0.18% | - **0.05%** (5.1 per 10,000) live birth incidence in England, using data from the NNRD in 2012.^28^ |

^†^ Neurodisability subgroups are non-mutually exclusive. Intellectual disability refers to cases defined using the primary case definition, including explicit diagnoses, high-risk conditions, and associated conditions; **^‡^** All cohort prevalence estimates are derived from HES data unless otherwise stated; ^§^ Published comparators vary in data source and ascertainment method.

CPRD: Clinical Practice Research Datalink; EUROCAT: European network of population-based registries for the epidemiological surveillance of congenital anomalies; HES: Hospital Episode Statistics; NCARDRS: National Congenital Anomaly and Rare Disease Registration Service; NDSCR: The National Down Syndrome Cytogenetic Register; NISALD: Northern Ireland Survey on Activity Limitation and Disability; NNRD: National Neonatal Research Database; NPD: National Pupil Database; SEND: Special Educational Needs and Disabilities

**Table S5. Cumulative mortality risk between ages 11 and 22 years among young people with intellectual disability or autism by ascertainment source.**

|  | **Girls** | | |  | **Boys** | | |
| --- | --- | --- | --- | --- | --- | --- | --- |
| **Neurodisability subgroup^‡^** | **N  (%)^a^** | **Deaths  (%)^b^** | **Cumulative risk  (95% CI)** |  | **N  (%)^a^** | **Deaths  (%)^b^** | **Cumulative risk  (95% CI)** |
| Intellectual disability (HES or NPD) | 17,143 | 397 | 3.1% (2.7-3.5%) |  | 29,680 | 533 | 2.6% (2.3-2.9%) |
| HES only | 6,402  (37%) | 54  (14%) | 1.3% (0.88-1.7%) |  | 8,466  (29%) | 81  (15%) | 1.6% (1.2-2.0%) |
| NPD only | 6,883  (40%) | 62  (16%) | 1.1% (0.81-1.4%) |  | 16,028  (54%) | 115  (22%) | 1.1% (0.83-1.3%) |
| HES and NPD | 3,858  (23%) | 281  (71%) | 10.1% (8.5-11.6%) |  | 5,186  (17%) | 337  (63%) | 9.3% (8.1-10.5%) |
| Autism (HES or NPD) | 7,744 | 24 | 0.52% (0.23-0.82%) |  | 43,307 | 106 | 0.38% (0.29-0.46%) |
| HES only | 755 (10%) | ^*^ | ^*^ |  | 2,724  (6.3%) | 56 (53%) | 1.3% (0.69-1.9%) |
| NPD only | 5,642 (73%) | 10  (41%) | 0.26% (0.07-0.45%) |  | 33,397  (77%) | 25  (24%) | 0.28% (0.19-0.37%) |
| HES and NPD | 1,347 (17%) | ^*^ | ^*^ |  | 7,186  (17%) | 25 (24%) | 0.51% (0.29-0.73%) |

^a^ n (% of total group); ^b^ n (% of total deaths); ^*^ Counts are suppressed where n < 10 to protect confidentiality and corresponding estimates are not reported; **^‡^** Neurodisability subgroups are non-mutually exclusive. Intellectual disability refers to cases defined using the primary case definition, including explicit diagnoses, high-risk conditions, and associated conditions.

CI: Confidence Interval; HES: Hospital Episode Statistics; NPD: National Pupil Database.

**S6 Table. Cumulative mortality risk between ages 11 and 22 years among young people with intellectual disability by case definition.**

|  | **Girls** | | |  | **Boys** | | |
| --- | --- | --- | --- | --- | --- | --- | --- |
| **Intellectual disability case definition** | **N** | **Deaths** | **Cumulative risk (95% CI)** |  | **N** | **Deaths** | **Cumulative risk (95% CI)** |
| Explicit diagnosis^†^ | 935 | 79 | 10.9% (7.7-14.0%) |  | 1478 | 80 | 7.3% (5.4-9.1%) |
| High-risk conditions**^‡^** | 2636 | 69 | 3.7% (2.5-5.0%) |  | 3022 | 59 | 2.9% (2.1-3.7%) |
| Associated conditions^§^ | 6685 | 187 | 4.1% (3.3-4.8%) |  | 9148 | 278 | 4.7% (4.0-5.3%) |

^†^ ICD-10 codes F70-73 and F78-79; **^‡^** Conditions with >75% probability of having intellectual disability; ^§^ Conditions with 30-75% probability of having intellectual disability.

CI: Confidence Interval; ICD-10: International Classification of Diseases 10^th^ revision.
